# Identification of loci genetically associated with antibodies to specific sets of citrullinated peptides in rheumatoid arthritis patients

**DOI:** 10.1101/2021.07.15.21259135

**Authors:** K Shchetynsky, B Brynedal, D Ramsköld, L Israelsson, M Hansson, L Mathsson-Alm, J Rönnelid, Rheumatoid Arthritis Consortium International (RACI), S Viatte, R Holmdahl, A Catrina, L Alfredsson, V Malmström, L Klareskog, D Gomez-Cabrero, L Padyukov

## Abstract

**Objectives:** Production of anti-citrullinated peptide/protein antibodies (ACPA) is characteristic for rheumatoid arthritis (RA) and may inform about biological pathways involved in disease development in specific subgroups. Since multiple loci in genome wide association screens have been implicated in RA risk, we investigated the association between genetic variations and the development of multiple types of ACPA in three big cohorts of RA patients from Sweden, USA and UK (EIRA, NARAC and WTCCC) with overall 6,127 individuals with RA.

**Methods:** We combined genotyping data from Illumina Immunochip with serological data for 16 ACPA specificities from a custom-made multiplex microarray (Thermo Fisher Scientific, ImmunoDiagnostic division). A logistic regression-based association test in each cohort was followed by meta-analysis.

**Results:** We report several loci, harbouring three or more variants associated with ACPA, reaching study-wide significance (FDR<0.1) – PTPN22, LINC02341_TNFSF11, THADA and TMEM174. Most of these loci are not known to be associated with RA. Stratification by “shared epitope” (SE) alleles indicated an association between the PTPN22 locus and levels of antibodies binding to the vimentin peptide, Cit-Vim60-75, in SE positive cases, but not in SE negative cases.

**Conclusion:** Our data identifies several new loci which associate with subsets of RA characterized by presence of specific ACPA and indicate unknown disease heterogeneity.

**Key messages:** *What is already known about this subject?:*

- ACPA define two subgroups within RA, and has been an important biomarker included in diagnostic criteria
- Commonly used ACPA test, anti-CCP, accumulate signals from autoantibodies to several peptides and does not discriminate between multiple autoantibody specificities.

*What does this study add?:*

- Our study demonstrates that different sets of ACPAs in RA have distinct genetic associations. Associated variants from our study localize in known regulatory regions and have transcriptomic effects beyond the associated locus
- We find that PTPN22 locus can drive positivity for multiple APCAs, while other found loci associate with specific autoantibody positivities

*How might this impact on clinical practice?:*

- Our study will extend personalized approach in handling of rheumatoid arthritis with focus on serologically defined subgroups of the disease.

## Introduction

Genetic association studies of complex diseases, including rheumatoid arthritis (RA), have helped discover multiple disease-associated loci in the human genome. Currently, more than 100 loci were implicated in RA susceptibility based on studies in multiple human populations[1]. It remains unclear how these genetic associations translate into clinically relevant phenotypes.

An increase of antibodies binding citrullinated proteins and peptides (ACPA) is characteristic for the major subset of RA. These antibodies were shown to bind to citrullinated filaggrin, from which a peptide-based assay, cyclic citrullinated peptide 1, was developed. It was succeeded by the anti-CCP2 assay that has been extensively used and studied for many years. A spectrum of ACPA was analysed for association with genetic variants of HLA locus recently[2]. However, the information regarding potential genetic associations of these RA-specific serotypes outside of the HLA locus is still lacking.

Identification of disease sub-traits has been useful for understanding genetic regulation of physiological pathways for several complex diseases[3, 4]. The high heterogeneity of complex diseases, including RA, suggests the existence of disease subgroups with distinct pathogenesis that potentially require different approaches to therapy. Earlier, we have found significant differences in genetic association between anti-CCP2 positive and anti-CCP2 negative RA[5, 6]. It is now commonly accepted that these two disease subgroups have different disease courses and clinical outcomes.

Current serological tests increased the granularity of RA-specific autoantibody analyses, facilitating quantification of multiple types of ACPA. Here, we integrate serological data with the Immunochip (a customized array designed to genotype ca 200,000 polymorphisms within 186 genetic loci associated with different autoimmune diseases) to detect RA subgroups[7]. We addressed association of 16 different autoantibodies with genetic variations in known immune-related loci. Due to predominately autoantibody negative status of controls genetic associations with specific serological markers may be difficult to discriminate from RA risk associations. We opted to use a case-only study design to address this limitation. To increase the power and to limit selection bias, we performed a meta-analysis including three independent cohorts with harmonized genetic and serological information. Here we focus on genetic associations with autoantibody specificities in RA cases that are found outside of the HLA locus. In contrast, the HLA-specific associations were covered in a separate study[2].

## Materials and methods

### Study cohorts

This study was performed within Rheumatoid Arthritis Consortium International (RACI)[8]. The case-only study includes 2,440 RA cases from the Epidemiological Investigation of Rheumatoid Arthritis (EIRA), 1,908 RA cases from North American Rheumatoid Arthritis Consortium (NARAC) and 2,040 RA cases from the Wellcome Trust Case Control Consortium (WTCCC). Data for case-control association was from our previous study[8]. Case-control expression analysis was based on data from the COMBINE study[9]. Table 1 represents the descriptive characteristics of the cohorts.

**Table 1.**
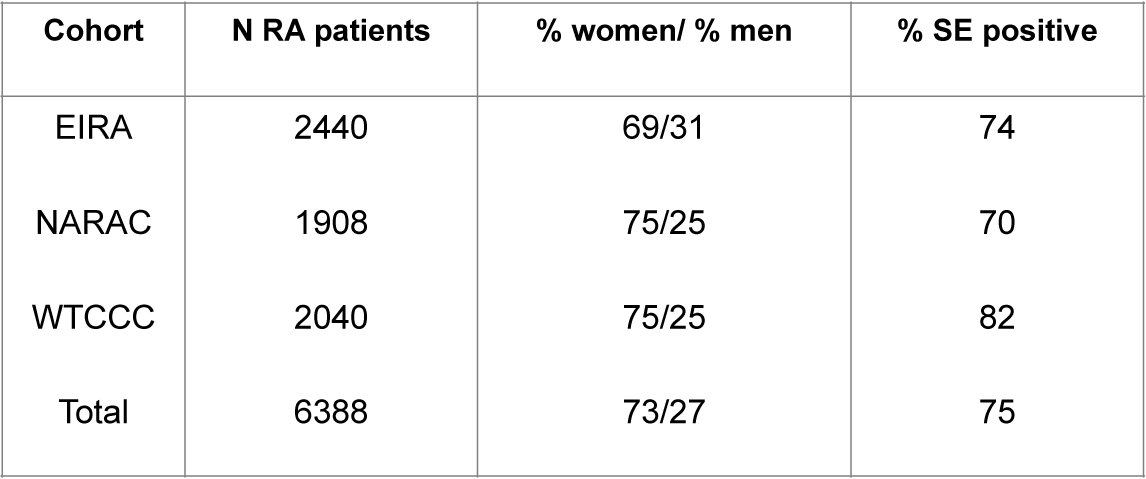
Description of the studied RA patient cohorts.

| **Cohort** | **N RA patients** | **% women/ % men** | **% SE positive** |
| --- | --- | --- | --- |
| EIRA | 2440 | 69/31 | 74 |
| NARAC | 1908 | 75/25 | 70 |
| WTCCC | 2040 | 75/25 | 82 |
| Total | 6388 | 73/27 | 75 |

Informed consent was obtained from all participants in all cohorts, in compliance with the latest version of the Helsinki Declaration. Regional ethical committees at correspondent sites have approved the study.

### Genotyping

Immunochip Illumina array data for all participating individuals was collected as previously described[8]. Amino acids defining the shared epitope (SE) allele status of patients were imputed from Immunochip data using SNP2HLA software[10]. Carriers of any number of alleles corresponding to DRB1*04; *01 (excluding *01:03) and *10 were regarded as SE-positive in the study. The attribution of imputed amino acid positions to HLA-DRB1 classes was performed according to criteria listed in Supplementary Table 1 (A, B, C). For samples from EIRA study the results of the imputation were compared to the HLA-DR typing with sequence-specific primer-PCR method (DR low-resolution kit; Olerup SSP, Saltsjöbaden, Sweden). The discrepancy between the imputation-based attribution and Olerup SSP method was below 2.5%. The algorithm used for inference of SE allele status from SNP2HLA output is available as a supplementary interactive MS Excel spreadsheet (Supplementary spreadsheet 1).

### Quality control of genetic data

In each of the 3 data sets of the discovery phase we applied the same quality check (QC) steps for the 22 autosomal chromosomes 1) removal of SNPs with missingness rate ≥0.01; 2) removal of individuals with missingness rate ≥0.05; removal SNPs that had differential missingness between cases and controls (p-value <0.001); 3) removal of SNPs that violated Hardy-Weinberg equilibrium (HWE) using a MAF-stratified p-value cut-offs according to the following criteria: MAF>0.3: p-value<0.0001, MAF 0.2 to 0.3: p-value<0.00001, MAF 0.1 to 0.2: p-value<0.000001, MAF<0.1: p-value<0.000001. 4) removal of SNPs with MAF <1%.

Prior to estimating the degree of relatedness between individuals, inbreeding coefficient and identity by descent we LD-pruned the autosomal chromosomes with the following settings in PLINK:

--indep-pairwise 100 2 0.1 (100 SNPs window, 2 SNPs step, r2=0.1). Next, we removed samples with inbreeding coefficient F≥0.05 or ≤ −0.05 and calculated identity-by-descent (IBD) estimates, quantified as pi-hat (proportion of IBD), excluding samples with values ≥0.1 (3rd-4th degree relatives). We used flashpca2 to calculate principal components (PCs) and removed individuals that were outside ± 6 standard deviations in each of the 10 first PCs. We further projected the samples of each data set onto the HapMap3 populations. Consensus SNPs were pulled from 3 cohorts, with addition of seven HapMap, release 3, cohorts – CEU, ASW, CHB, JPT, TSI, GIH, YRI. We manually removed any individuals from RA cohorts that fell outside HapMap CEU and the main analysis cluster (Figure 1B). Overall, out of 6,388 individuals and 200,243 variants, 6,127 RA patients and 94,146 variants have cleared these QC steps and were included in subsequent association analysis.

**Figure 1.**
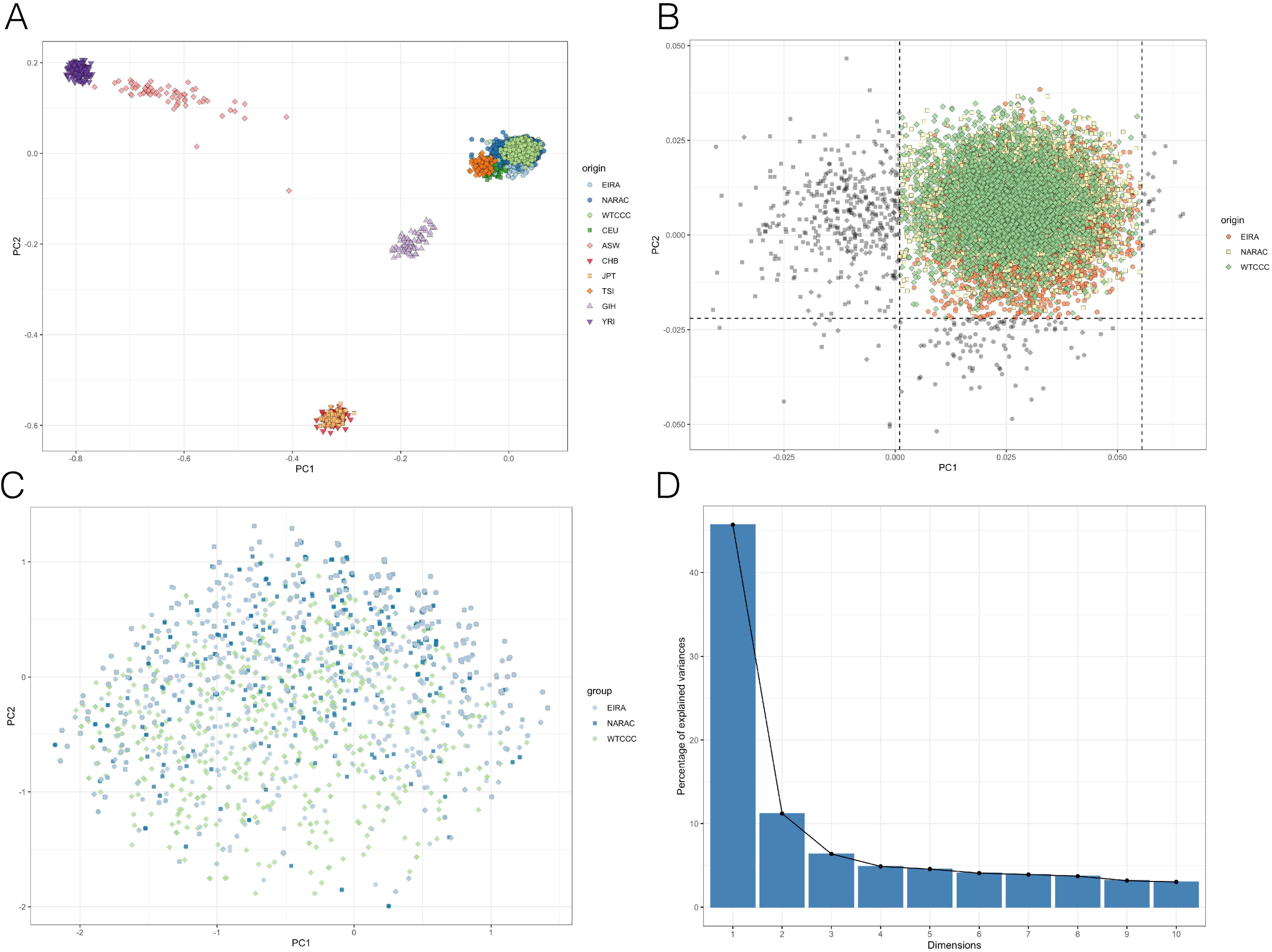
**A)** PCA of the 3 RA cohorts projected on HapMap3 data. **B)** Manual curation of the study cluster based on the first 2 principal components of genetic variant PCA of EIRA, NARAC and WTCCC cohorts. Individuals with grey symbols were removed from analyses. **C)** PCA based on data for 16 autoantibody specificities from the study cohorts. **D)** Percentage of variance explained by the serological phenotype of RA patients from the 3 study cohorts.

### Serological data

The serological profiles were generated with the help of a custom-made multiplex microarray (Thermo Fisher Scientific, ImmunoDiagnostics). Final profiling in the current study included 16 RA related citrullinated peptides and their native arginine-containing counterparts (Supplementary Table S1)[11-21]. Sera from 551 population-based controls matched for age and sex were analysed to determine the cutoff levels of positive reactions for each ACPA specificity corresponding to 98% specificity of the test. A full description of this technology, with extensive validation of the chip-based technique in comparison with ELISA-based technology, and a review of the diagnostic performance of this microarray system was published elsewhere[13, 22]. Each serological phenotype was coded as a binary category for further analyses.

### RNA-sequencing

RNA-seq data from 92 newly-diagnosed RA patients prior to first treatment and from 52 patients out of this group 3 months after treatment with methotrexate (totally 144 RNA samples) were obtained as part of COMBINE project[9]. RNA was isolated from PBMC samples separated from citrated blood using Ficoll-Paque (GE Healthcare, Sweden). It was sequenced using an Illumina HiSeq 2000, the TruSeq RNA sample preparation kit and 2×100bp paired-end setup resulting in mean read depth 15.7M read-pairs per sample. The alignment was performed using Tophat2[23] followed by “number of reads per gene” count matrix generation using htseq-count[24].

### Statistics

Principal component analysis (PCA) of serological and genetic features was performed using R princomp package and flashpca2, respectively. Immunochip-based PCA was done as described[25]. Association analyses and meta-analyses were performed with PLINK 1.9 using logistic regression with adjustment for sex and first 5 principal components for genetic data. The FDR<0.1 cutoff was used to call significant associations. Loci containing three or more SNPs with FDR<0.1 were assigned as credible associations. The differential expression analysis between genotype groups was carried out on raw counts by DESeq2 package[26] using regression analysis model *expression ∼ genotype * treatment* to account for the potential effects of methotrexate treatment on gene expression.

## Results

### Cohort homogeneity

First, the Immunochip data was pruned for genotype missingness, SNPs with low minor allele frequency and 12,574 SNPs from 4 high LD regions were removed as suggested previously[25]. We restricted the study cohort to the individuals that clustered with CEU samples (Figure 1A).

Second, we tested our cohorts for autoantibody-related phenotypic variability. To verify homogeneity between cohorts, we performed PCA across autoantibody positivity data for 16 specificities in the three combined cohorts. This analysis did not show any cohort-specific contribution to the variance, suggesting similar distribution of the phenotypes for different cohorts (Figure 1C). The first 2 components accounted for 50% of the variance with a sharp decline for the other PCs (Figure 1D).

We have performed association analysis for each of the peptides in each of the 3 cohorts. The association summary statistics were combined in a meta-analysis (performed for each peptide).

### Correlation between different ACPA specificities

A certain level of correlation was expected between studied ACPA and we addressed it in a hierarchical clustering analysis for all three cohorts jointly (Supplementary figure 1). We identified at least 3 clusters of autoantibody co-occurrence. However, the correlation between autoantibodies is generally only modest (r^2^≤0.6).

### Genetic associations to anti-citrullinated peptide autoantibodies in rheumatoid arthritis

We performed case-only genetic association study involving 6,127 individuals and 94,146 genetic variants comparing autoantibody positive vs. autoantibody negative RA individuals for each of 16 ACPA specificities, first - independent for each study cohort and then - in meta-analysis. For simplicity, we present results from meta-analyses while data for individual cohorts are included in supplemental materials.

Overall, the meta-analysis of the 3 cohorts revealed 12 loci on several chromosomes associated with the presence of one or more ACPA (Figure 2). Out of these, we nominated four loci containing three or more variants associated with at least one of ACPA specificities at FDR p-value above 0.1 as credible association peaks. The four loci were PTPN22, LINC02341_TNFSF11, TMEM174 and THADA.

**Figure 2.**
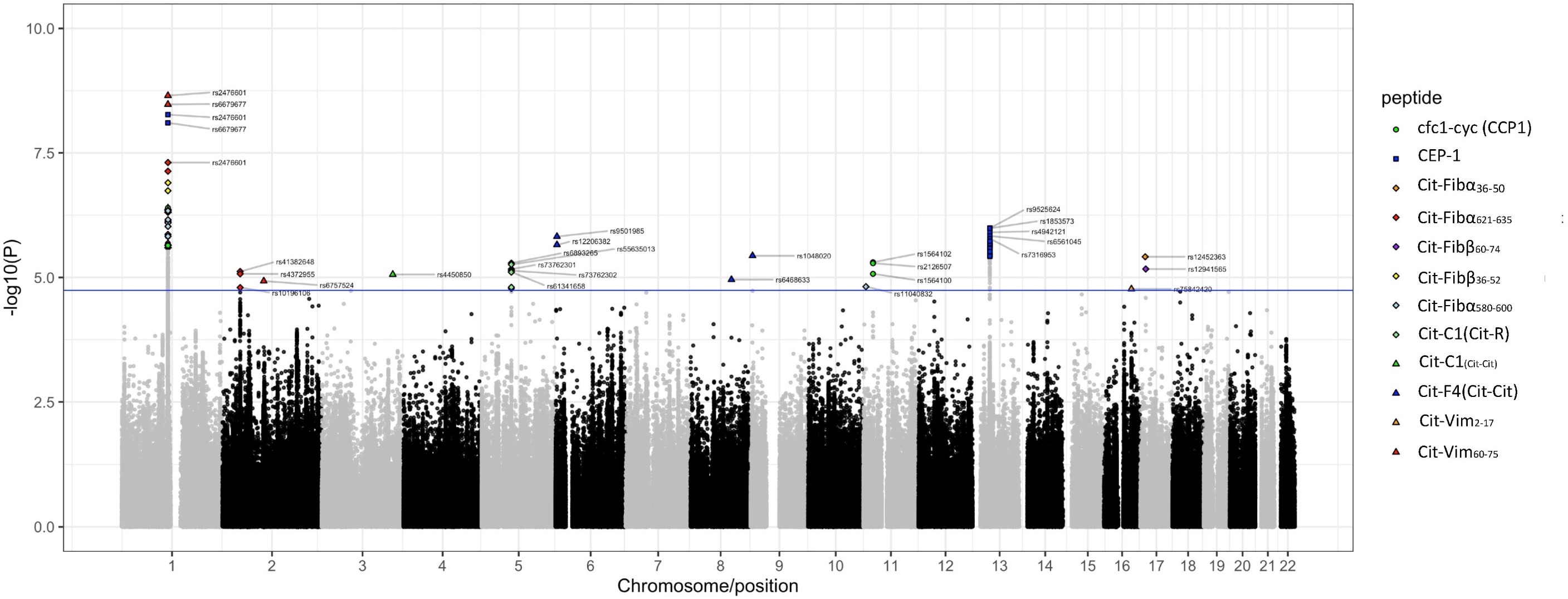
Composite plot for 16 auto antibody meta-analyses of association between 94,146 Immunochip variants for 6,127 RA cases from EIRA, NARAC and WTCCC. Solid line – FDR=0.1

Specifically, the PTPN22 rs2476601 from chromosome 1 was in significant association with 9 types of autoantibody specificities, including Cit-Vim_60-75_, CEP-1, Cit-Fibβ_36-52_ (p-value range 1.57×10^−5^-2.24×10^−9^; OR range 1.19–1.28) (Supplementary Datasheet SD1). These associations were also observed for neighbouring SNP rs6679677, which is in full LD with SNP rs2476601. Locus THADA (chromosome 2) contained three variants associated with Cit-Fibα_621-635_ and one association to Cit-Vim_60-75_ ACPA specificities (p-value ranges 6.532 × 10^−6^ – 1. 1 6 3 × 1 0 ^-5^; OR ranges 1. 1 8 – 1. 2 8). In contrast, LINC02341_TNFSF11 locus contained 59 SNPs associated with a single CEP-1 ACPA specificity (p-value range 2.436×10^−7^-9.567×10^−6^; OR range 1.17–1.25) (Supplementary Datasheet SD1). Similarly, the locus TMEM174 (chromosome 5) contained seven associated variants to just one ACPA specificity – Cit-C1_(Cit-R)_.

Additionally, we detect several isolated association signals for SNPs on chromosomes 3, 8, 9, 19 for various ACPA specificities that did not reach our criteria for a credible associated locus (Supplementary Datasheet SD1).

### Regulatory context of associated variants

To provide explanation for functional background of found associations we assessed if the associated variants perturbed known regulatory elements of the genome (Supplementary Table 2) and found that these variants overlap with a number of CCCTC-binding factor (CTCF) binding sites, promoter regions, and transcription-factor (TF) binding sites. Specifically, rs17013326 from the PTPN22 locus perturbs a known CTCF binding site, while variants rs61817589, rs11552449, rs11552449, rs3811019 affect known promoter regions. Variants rs12870516 and rs2062305 from the LINC02341_TNFSF11 locus perturb 2 separate CTCF binding sites, while variants rs12871509, rs9594738 and rs56008941 are located in known transcription factor binding sites. Variants rs41382648 and rs10196106 from the THADA locus overlap known enhancer regions.

### Serotype-genotype associations in RA subgroups defined by HLA-DRB1 SE allelic group

To isolate potential influence of *HLA-DRB1* SE alleles on non-HLA serotype-genotype associations in the context of previous observations we performed two separate meta-analyses of RA patients stratified by presence of *HLA-DRB1* SE alleles. In the isolated meta-analysis of 4,832 *HLA-DRB1* SE allele positive RA patients from 3 cohorts the association of rs2476601 and rs6679667 to Cit-Vim_67-85_ remained highly significant (p-value range 5.352×10^−7^ — 7.567×10^−6^; FDR<0.05) (Supplementary Figure 2A). Association hits on chromosomes 6 and 13 could also be seen although after decreasing of the number of individuals in meta-analysis both did not pass the 0.1 FDR cutoff. However, neither these nor any other associations between serological markers and genotype could be observed in the meta-analysis of 1,556 *HLA-DRB1* SE allele negative patients (Supplementary Figure 2B). Minor allelic frequencies for SNPs associated with ACPA presented separately for *HLA-DRB1* SE allele positive and negative patient’s groups in Supplementary Table S4.

### Comparison with a case-control study

We performed a case-control analysis in 3 independent steps: all controls vs. autoantibody positive RA patients, all controls vs. autoantibody negative RA patients, and all controls vs. all RA patients. In these analyses association was driven by genetic association with RA, and in all 3 analyses the association landscape was markedly different to the results of our case-case study, but was largely consistent between these 3 analyses (Supplementary Figure 4A, B and C respectively).

### Effects of ACPA-associated variants on transcriptome

Most regions with associations to autoantibody demonstrated a high level of LD. Based on the highest value of linkage (r^2), we nominated 7 tagging SNPs (rs10858000, rs4372955, rs4450850, rs9379881, rs1048020, rs12871509 and rs504850) representing the serologically associate loci from chromosomes 1, 2, 5, 6, 9, 13 and 19. We assessed the effects of chosen tagging SNPs on global transcription in PBMCs from a group of 92 newly diagnosed RA patients. 75 genes were found differentially expressed with fold changes above 2 and FDR <0.05 based on genotypes of serotype-associated SNPs. The majority of differentially expressed (DE) genes (n=46) were affected by tagging SNP from the PTPN22 locus at chromosome 1 (Figure 3 A).

**Figure 3.**
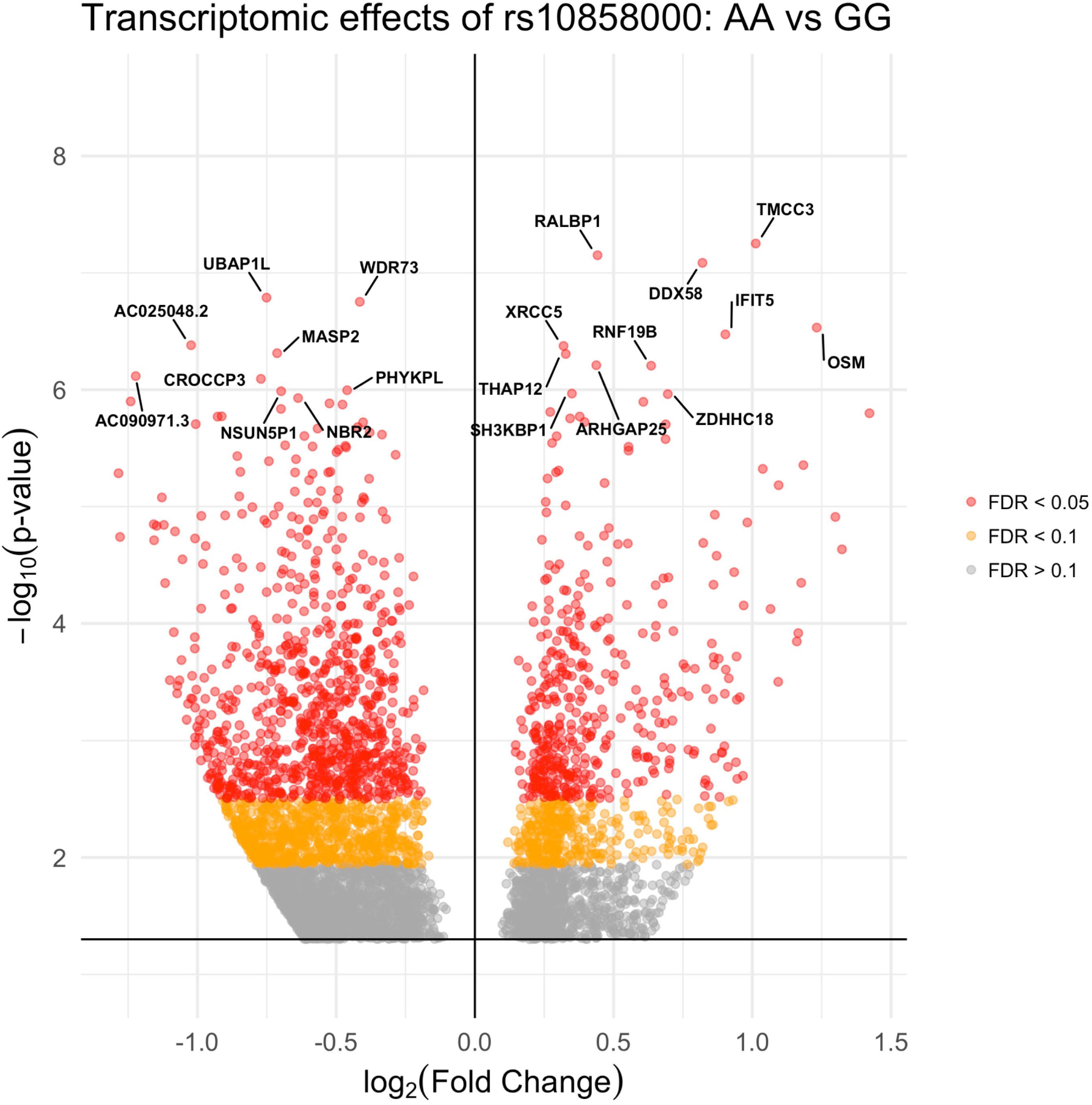
Influence of tagging autoantibody specificity associated variants on gene expression. Transcriptional consequences of AA vs GG homozygote of the tagging rs10858000 variant from chromosome 1 (PTPN22 locus).

## Discussion

Our investigation of 6,127 individuals with RA for genetic association with ACPA specificities demonstrates several association signals at PTPN22, LINC02341_TNFSF11, TMEM174 and THADA loci. We show that *PTPN22* SNP rs2476601 is associated with the presence of multiple autoantibodies. This polymorphism was established as the second strongest risk factor for RA after *HLA-DRB1* SE in multiple studies[27-29]. It was shown to be associated with ACPA-positivity in RA, as defined by a general anti-CCP test. Our data indicate that the involvement of rs2476601 in RA is not specific for an isolated serological subtype of the disease, but it is generally associated with ACPA-positivity in RA. We found that 9 out of the 16 ACPA specificities assessed were associated with this locus in comparison to RA patients without autoantibodies. Our current analysis confirms the association of autoantibody positive RA with PTPN22 locus, however we were able to get a detailed view on that association, suggesting it is a composite measure for associations with multiple ACPA specificities, and demonstrating possible pleiotropic effect regarding autoantibody phenotypes in RA. Conversely, most remaining associations were restricted to single ACPA specificities (Figure 4) and were not previously found in GWAS for RA. It is indicative that in our study we detected associations between genetic and serological markers regardless of the difference in frequency of specific serological phenotypes in the RA group (Suppl. Table 1).

**Figure 4.**
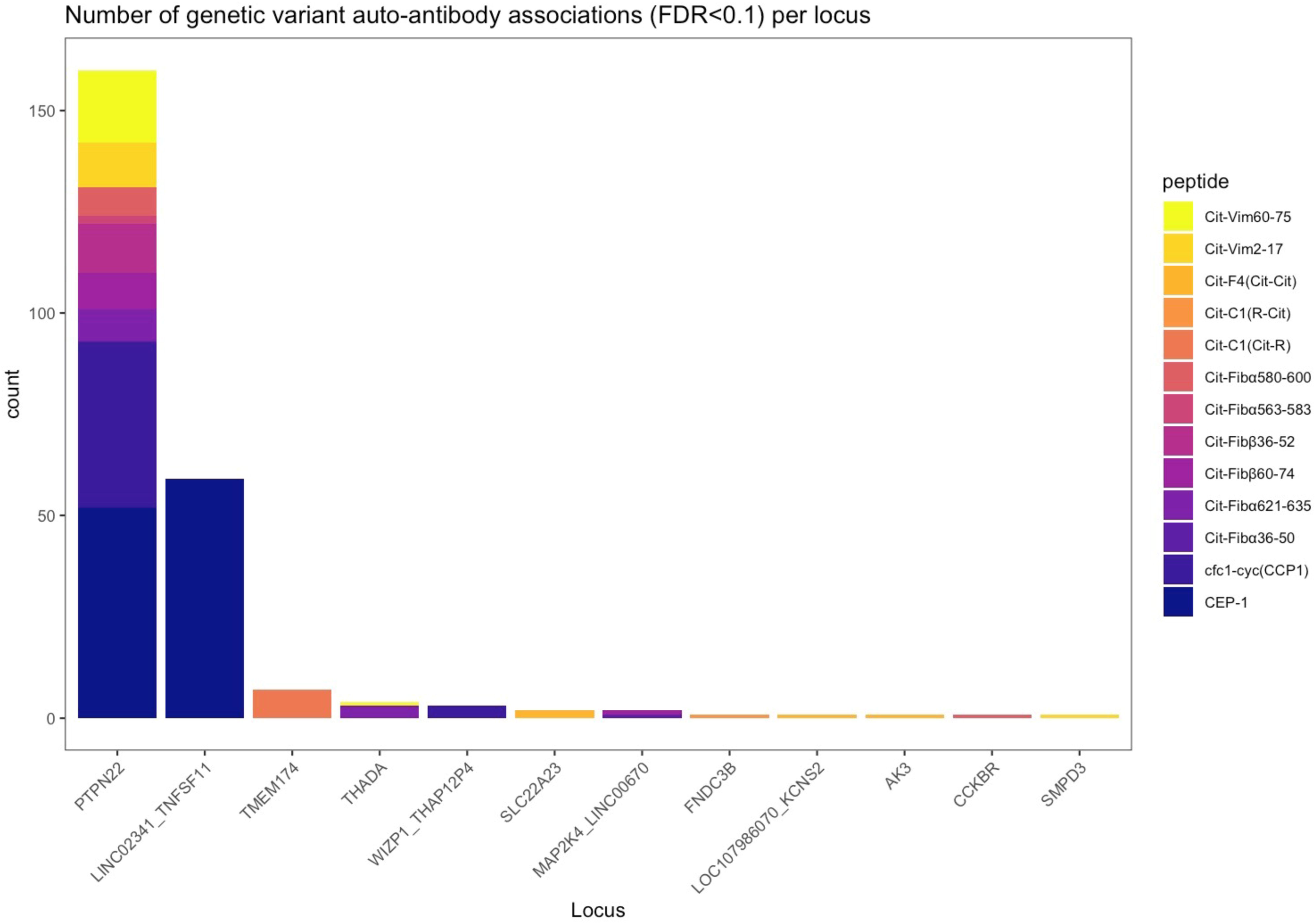
Number of variants associated to ACPA specificities per genetic locus. Credible loci were defined as 3 or more variants associated with any peptide with FDR p-value<0.1

In this study we show, for the first time, that LINC02341_TNFSF11 locus on chromosome 13 is associated with anti-CEP-1 positivity in RA patients. Associated variants are located in lncRNA LINC02341 locus and were reported as whole blood eQTLs for neighbouring genes *AKAP11* and *TNSF11* (*GTEx* project). This finding warrants further inquiries into functional connections between this locus and CEP-1 positive RA.

Additionally, serotype associations were found for variants in the gene THADA. This locus was found in association with multiple sclerosis, type 2 diabetes and polycystic ovary syndrome and was shown to be differentially regulated in pathogenic CD4+ T cells[30-32].

We have shown that several variants from the PTPN22, LINC02341_TNFSF11 and THADA loci perturb known genomic regulatory elements, including promoters enhancers and transcription-factor-binding sites (Table S2). Together, these findings support the utility of sub-phenotyping of RA based on ACPA specificities for genetic studies and suggest further studies in relation to clinically important phenotypes. Using this approach, we discovered at least three candidate loci that were not previously shown as pertinent to RA. Although we see that the effect size for these associations is relatively modest, our meta-analysis provides sufficient power to detect these effects.

Generally, genetic associations with specific ACPA-associated markers are difficult to discriminate from RA risk associations due to predominately autoantibody negative status of controls, prompting us to adopt a case-only study design. This study aims at understanding patient heterogeneity within RA, rather than addressing susceptibility to RA with a certain serological profile.

In our study, no significant associations from the general meta-analysis were observable in the *HLA-DRB1* SE-negative subgroup of RA cases, whereas some remained significant in the *HLA-DRB1* SE-positive subset. The association of *PTPN22* rs2476601 with several ACPA in *HLA-DRB1* SE positive RA is consistent with previous findings[33]. Our data suggest that the strongest discovered associations with certain ACPA specificities in RA on chromosomes 1, 6 and 13 may be exclusive for patients positive for *HLA-DRB1* SE. However, it is possible that the signals are abated by lower power in *HLA-DRB1* SE-negative patients, which represent a minority of RA cases.

This study contributes to our understanding of genetic variation contributing to the ACPA positivity in RA, suggesting that the genetic variant association to ACPA may vary depending on which specificities are present. Notably, however, the cross-reactivity of ACPA with different citrullinated peptides and proteins is extensive[34, 35]. Our study suggests that development of ACPA with a certain profile of specificities is unlikely to be a random process, since the number of specificities associated with genetic loci is more than expected by chance. These associations may suggest a modifying effect of reported genetic loci on the risk of ACPA positive RA.

Among the study’s weaknesses is the heterogeneity of RA patients from three cohorts with different selection criteria. This restricts us to detecting only major effects that are not masked by heterogeneity. Secondly, our study utilised Illumina Immunochip data, limited to known immune-related loci by design. A genome-wide dataset may reveal more genetic associations with antibodies against specific ACPA in RA.

In summary, we have identified associations between serological sub-phenotypes in RA and multiple genetic loci, most of which were not implicated in the disease originally. While previously reported PTPN22 locus contributes broadly to a spectrum of ACPA specificities, we find other loci mostly associated to the presence of individual autoantibodies in RA

## Supporting information

Supplementarty Spread Sheet 1

Supplementary Data Set 1

## Data Availability

Data are available upon reasonable request

## Contributorship

Design of the project: KS, BB, VM, DGC, LP. Data generation: Data analysis: Methodological contribution: Manuscript writing: Manuscript review: all.

## Acknowledgements

We like to thank members of RACI, EIRA, NARAC and WTCCC for giving access to genotyping data and for collaboration. KS was supported by Börje Dahlin’s foundation. LP is supported by the Swedish Research Council.

## Funding, grant/award info

Vetenskapsrådet (2015-3006, 2018-2399, 2018-2884), Reumatikerförbundet, Stiftelsen Börje Dahlins Fond, Stiftelsen Jubileumsklinikens Forskningsfond.

## Ethical approval information

Stockholm Regional Ethics Committee: Dnr 96-174, 02-288, 210/935-31/1, 2021-00125.

## Data sharing statement

Data are available upon reasonable request from Dr Leonid Padyukov.

## Patient and Public Involvement

Not applicable.

