## Supplementary Figures for "Identification of loci genetically associated with antibodies to specific sets of citrullinated peptides in rheumatoid arthritis patients"

Supplementary Figure 1

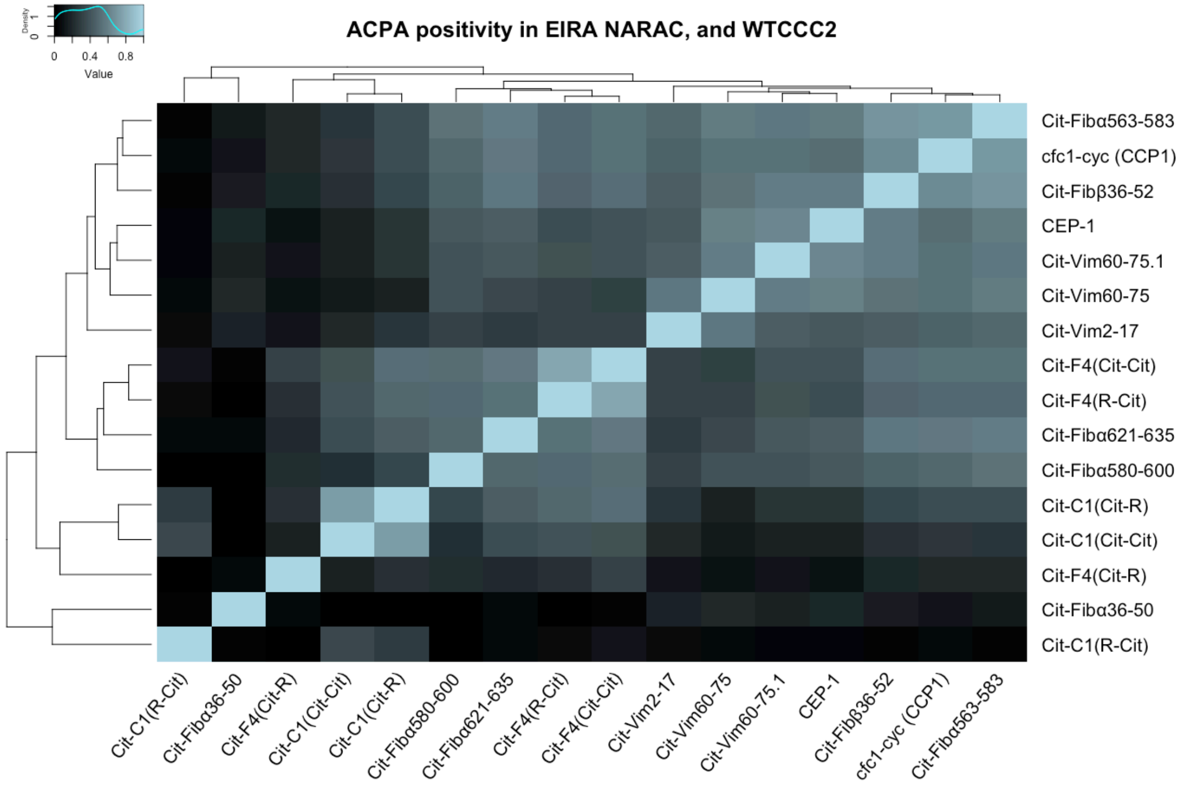


Correlation of of positivity for each of the 16 studied ACPA specificities in 3 cohorts from Sweden.

Supplementary Figure 2

**A**

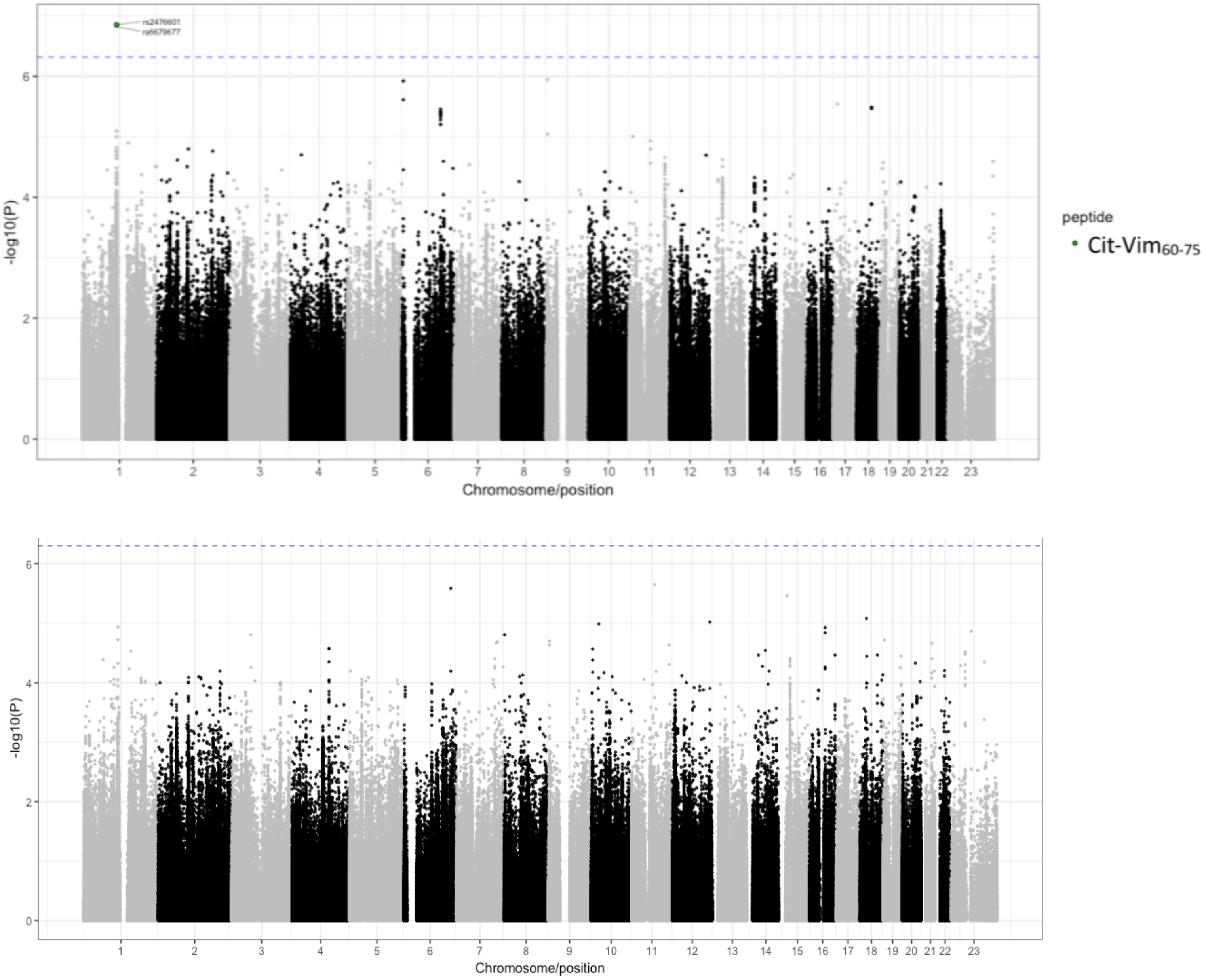


Meta-analysis of genetic association to 16 auto-antibody specificities in EIRA, NARAC and WTCCC cohorts stratified by the presence of shared epitope.

Meta-analysis of SE-positive patients

Meta analysis of SE-negative patients

Dashed line — FDR corrected p-value of 0.05

Supplementary Figure 3

**A**

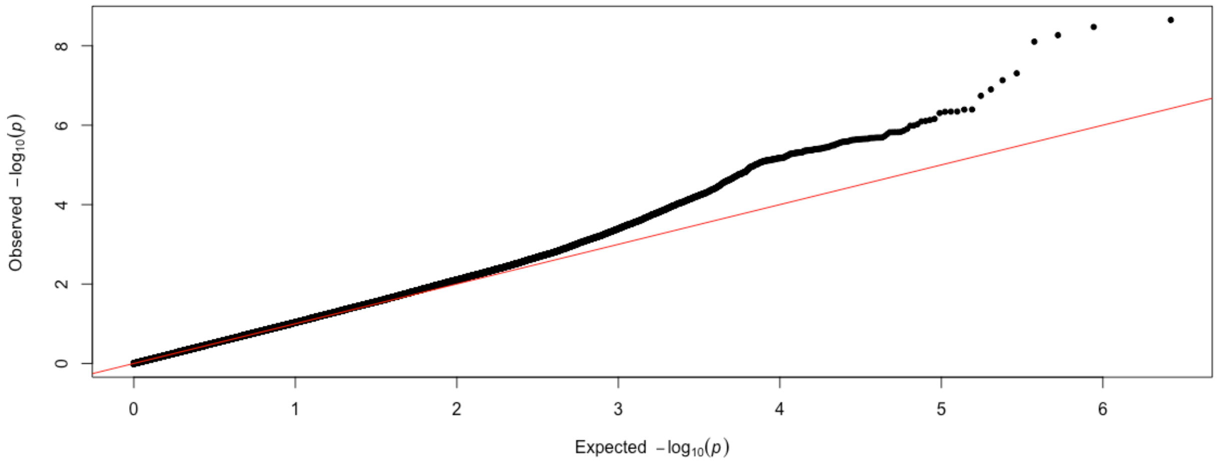


**B**

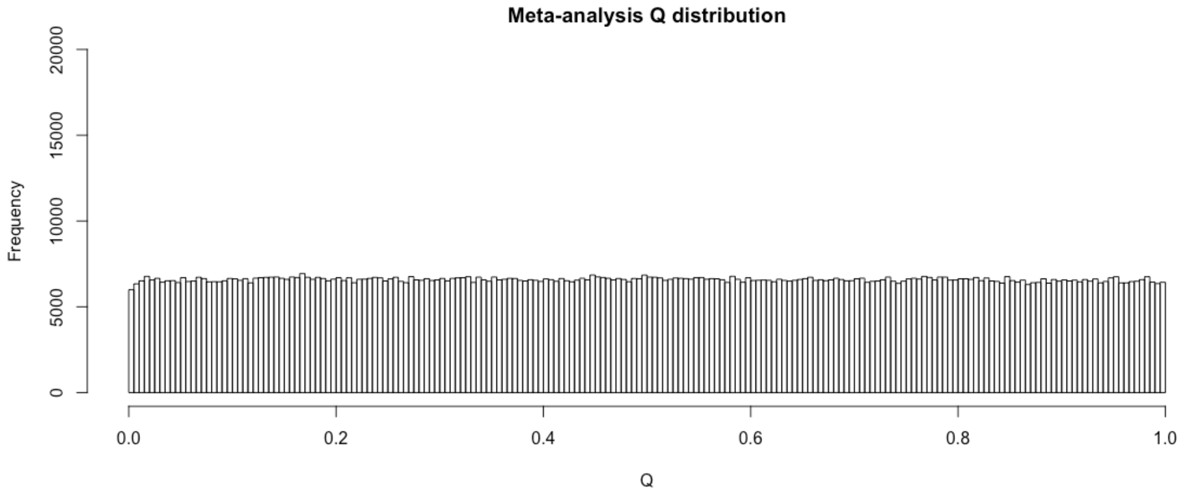


The quality metrics of the meta-analysis of genetic association to specific RA auto-antibody positivity. A) The observed vs expected distribution of test p-values for each marker (all peptide associations)

B) The Cohran's Q distribution for each marker (all peptide associations)

Supplementary Figure 4

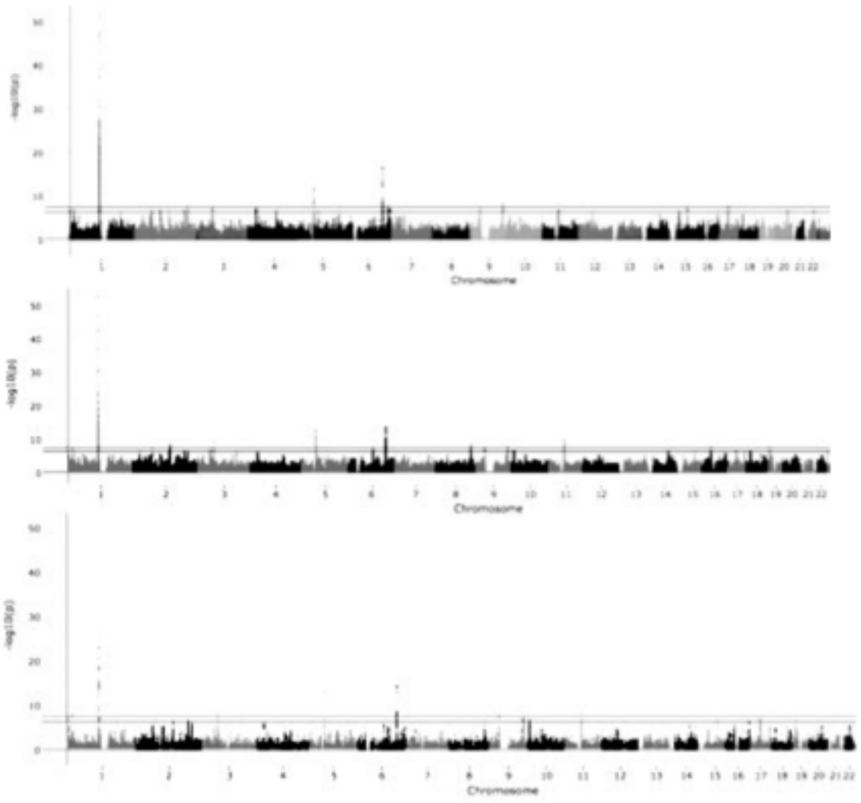

Case-control meta-analysis of the EIRA, NARAC and WTCCC cohorts.

A) Healthy controls vs auto-antibody positive RA patients. B) Healthy controls vs auto-antibody negative RA patients. C) Healthy controls vs all RA patients

Solid red line — FDR corrected p-value of 0.02
Dashed blue line — FDR corrected p-value of 0.05
