## Supplementary Tables for "Identification of loci genetically associated with antibodies to specific sets of citrullinated peptides in rheumatoid arthritis patients"

**Table S1. Prevalence of inidividual auto-antibody specificities across RA patients in EIRA, NARAC and WTCCC cohorts using Phadia cip assay**

| Peptide name (Reed et al.) | Protein | % positive EIRA | % positive NARAC | % positive WTCCC |
|---|---|---|---|---|
| Cit-Fibα$_{563\text{-}583}$ | Fibrinogen alpha chain | 43.61 | 53.88 | 52.40 |
| Cit-Vim$_{60\text{-}75}$ | Vimentin | 45.98 | 58.81 | 59.36 |
| cfc1-cyc (CCP1) | Filaggrin | 32.30 | 51.36 | 38.53 |
| Cit-Vim$_{2\text{-}17}$ | Vimentin | 47.38 | 48.64 | 55.20 |
| CEP-1 | alpha-enolase | 41.48 | 50.16 | 46.37 |
| Cit-Fibα$_{621\text{-}635}$ | Fibrinogen alpha chain | 25.08 | 45.39 | 32.25 |
| Cit-Fibα$_{580\text{-}600}$ | Fibrinogen alpha chain | 47.75 | 55.61 | 70.83 |
| Cit-Fibβ$_{36\text{-}52}$ | Fibrinogen beta chain | 14.18 | 13.47 | 34.75 |
| Cit-Fibα$_{36\text{-}50}$ | Fibrinogen alpha chain | 11.11 | 9.91 | 25.93 |
| Cit-Fibβ$_{60\text{-}74}$ | Fibrinogen beta chain | 4.59 | 2.46 | 15.39 |
| Cit-C1$_{(Cit\text{-}R)}$ | type-II collagen | 24.80 | 23.48 | 52.40 |
| Cit-C1$_{(Cit\text{-}Cit)}$ | type-II collagen | 27.91 | 27.62 | 48.38 |
| Cit-C1$_{(R\text{-}Cit)}$ | type-II collagen | 6.72 | 11.48 | 29.56 |
| Cit-F4$_{(R\text{-}Cit)}$ | type-II collagen | 34.59 | 28.25 | 51.47 |
| Cit-F4$_{(Cit\text{-}Cit)}$ | type-II collagen | 17.21 | 13.16 | 15.20 |
| Cit-F4$_{(Cit\text{-}R)}$ | type-II collagen | 58.36 | 64.57 | 28.14 |

**Table S2. Genomic regulatory eliments perterbed by AB-associated SNPs (hg19)**

| SNP | Location | Allele | Feature | Regulatory element |
|---|---|---|---|---|
| rs17013326 | 1:113546694-113546694 | A | ENSR00000253653 | CTCF_binding_site |
| rs61817589 | 1:113758713-113758713 | T | ENSR00000011611 | promoter |
| rs11552449 | 1:113905767-113905767 | G | ENSR00000011633 | promoter |
| rs11552449 | 1:113905767-113905767 | T | ENSR00000011633 | promoter |
| rs3811019 | 1:113928961-113928961 | G | ENSR00000011636 | promoter |
| rs12871509 | 13:42377939-42377939 | C | ENSR00000061906 | TF_binding_site |
| rs9594738 | 13:42378009-42378009 | T | ENSR00000061906 | TF_binding_site |
| rs56008941 | 13:42378475-42378475 | T | ENSR00000061907 | TF_binding_site |
| rs12870516 | 13:42381829-42381829 | C | ENSR00000271772 | CTCF_binding_site |
| rs2062305 | 13:42478744-42478744 | A | ENSR00000271780 | CTCF_binding_site |
| rs504850 | 19:55654109-55654109 | T | ENSR00000111697 | promoter |
| rs41382648 | 2:43305385-43305385 | T | ENSR00000116158 | enhancer |
| rs10196106 | 2:43344768-43344768 | T | ENSR00000116170 | enhancer |
| rs4450850 | 3:172171677-172171677 | C | ENSR00000161921 | enhancer |

**Table S3 A. Aminoacid positions determining DRB1:01**

| AA_DRB1_11_32660115_L | AA_DRB1_67_32659947_L | AA_DRB1_70_32659938_Q | AA_DRB1_71_32659935_R |
|---|---|---|---|
| AA | AP | AP | AA |
| AA | PP | PP | PA |
| PA | PP | PP | PA |
| PA | AP | AP | PA |
| PA | AP | AP | PP |
| AA | AP | AP | PA |
| PP | PP | PP | PP |
| AA | AA | AA | PA |
| AA | AP | PP | AA |
| AA | AA | AA | AA |
| AA | AP | PP | PA |
| PA | AP | PP | PA |
| AA | AA | AP | PA |
| AA | AA | AA | PP |
| AA | PP | PP | AA |
| AA | AA | AP | AA |
| AA | AA | PP | AA |
| AA | PP | AP | PA |
| AA | AP | AA | PP |
| AA | AP | AA | PA |
| AA | AP | AP | PP |
| PA | AA | AA | AA |
| PA | PP | PP | PP |
| PA | AA | AA | PA |
| AA | PP | AA | PP |
| PA | AP | AP | AA |
| PA | PP | AP | PP |
| PA | AP | AA | PA |
| AA | PP | AP | PP |
| AA | PP | PP | PP |
| PP | AP | AP | PA |
| PA | AA | AP | AA |
| PP | AA | AA | AA |
| PP | AP | PP | PA |

**Table S3 B. Aminoacid positions determining DRB1:10**

| AA_DRB1_9_32660121_E | AA_DRB1_10_32660118_E | AA_DRB1_11_32660115_V | AA_DRB1_30_32660058_R | AA_DRB1_31_32660055_V | AA_DRB1_32_32660052 |
|---|---|---|---|---|---|
| PP | AA | AA | AA | AA | HH |
| PP | AA | PP | AA | AA | YY |
| AP | AA | PA | AA | AA | YY |
| AP | AA | AA | AA | AA | HY |
| AA | AA | AA | AA | AA | YY |
| AP | PA | PA | PA | PA | HY |
| PP | AA | PA | AA | AA | YY |
| PP | AA | PA | AA | AA | HY |
| PP | AA | AA | AA | AA | HY |
| AP | AA | AA | AA | AA | YY |
| PP | PA | PA | PA | PA | HH |
| PP | AA | AA | AA | AA | YY |
| PP | PA | PP | PA | PA | HY |
| PP | PA | PA | PA | PA | HY |
| PP | PP | PP | PP | PP | HH |
| PP | AA | AA | PA | PA | HH |

**Table S3 C. Aminoacid positions determining DRB1:04**

| AA_DRB1_9_32660121_E | AA_DRB1_10_32660118_Q | AA_DRB1_11_32660115_V | AA_DRB1_26_32660070_F | AA_DRB1_28_32660064_D | AA_DRB1_31_32660055_F | AA_DRB1_33_32660049 |
|---|---|---|---|---|---|---|
| PP | AA | AA | AP | PP | PP | NN |
| PP |  | PP | PP | PP | PP | HH |
| AP | PP | PA | AP | AP | AP | HN |
| AP | AP | AA | AP | AP | AP | NN |
| AA | PP | AA | AP | AA | AP | NN |
| AP | AP | PA | AP | AP | AP | NN |
| PP | AP | PA | PP | PP | PP | HN |
| AA | PP | AA | AA | AA | AA | NN |
| AP | AP | AA | PP | AP | PP | NN |
| PP | AP | PA | AP | AP | PP | HN |
| AA | PP | AA | AP | AP | AP | NN |
| AP | AP | AA | AP | PP | PP | NN |
| PP | AA | AA | PP | PP | PP | NN |
| PP | AP | PA | AP | PP | PP | HN |
| AP | PP | PA | PP | PP | PP | HN |
| AA | PP | AA | PP | AP | PP | NN |
| PP | AA | AA | AA | AA | PP | NN |
| PP | AA | AA | AA | PP | PP | NN |
| AP | PP | PA | PP | AP | PP | HN |
| AP | AP | AA | PP | PP | PP | NN |
| AA | PP | AA | PP | PP | PP | NN |
| AA | PP | AA | PP | AA | PP | NN |
| AP | AP | AA | AP | AP | PP | NN |
| AP | AP | PA | AP | AA | AP | NN |
| AP | AP | AA | AA | AP | AP | NN |
| PP | AA | PA | AP | AP | AP | NN |
| AP | AP | AA | AA | AA | AP | NN |
| PP | AA | AA | AP | AP | PP | NN |
| AP | AP | PA | AA | AA | AA | NN |
| PP | AP | PA | PP | AP | PP | HN |
| PP | AA | AA | AA | AP | PP | NN |
| PP | AP | PP | AP | AP | AP | HN |
| PP | AA | PA | AA | AP | AP | NN |
| PP | AA | AA | AP | AA | PP | NN |
| AP | AP | AA | AP | AA | PP | NN |
| PP | AA | PP | AA | AA | AA | NN |
| PP | AA | AA | PP | AP | PP | NN |
| AP | AP | AA | AP | AA | AP | NN |
| PP | AA | AA | AP | PP | PP | HN |
