## Supplementarty Spread Sheet 1 for "Identification of loci genetically associated with antibodies to specific sets of citrullinated peptides in rheumatoid arthritis patients"

| FID | IID | GENDER | CASE | AA_DRB1_71_32659935_R | | AA_DRB1_70_32659938_Q | | AA_DRB1_67_32659947_L | | AA_DRB1_33_32660049 | | AA_DRB1_32_32660052 | | AA_DRB1_31_32660055_F | | AA_DRB1_31_32660055_V | | AA_DRB1_30_32660058_Y | | AA_DRB1_30_32660058_R | | AA_DRB1_28_32660064_D | | AA_DRB1_26_32660070_F | | AA_DRB1_11_32660115_V | | AA_DRB1_11_32660115_L | | AA_DRB1_10_32660118_Q | | AA_DRB1_10_32660118_E | | AA_DRB1_9_32660121_E | | CODE10 | CODE04 | CODE01 | IF10 | IF04 | IF01 | CALL |
|---|---|---|---|---|---|---|---|---|---|---|---|---|---|---|---|---|---|---|---|---|---|---|---|---|---|---|---|---|---|---|---|---|---|---|---|---|---|---|---|---|---|---|
| FID_Example1 | IID_Example1 | 2 | 1 | A | A | A | P | A | P | N | N | H | H | P | P | A | A | P | P | A | A | P | P | A | P | A | A | A | A | A | A | A | A | P | P | PP AA AA AA AA HH | PP AA AA AP PP PP NN | AA AP AP AA | 0 | 0 | 0 | N |
| FID_Example2 | IID_Example2 | 2 | 1 | P | A | P | P | P | P | H | H | Y | Y | P | P | A | A | P | P | A | A | P | P | P | P | P | P | A | A | P | P | A | A | P | P | PP AA PP AA AA YY | PP PP PP PP PP PP HH | AA PP PP PA | 0 | IS04 | 0 | SE |
| FID_Example3 | IID_Example3 | 2 | 1 | P | A | P | P | P | P | H | N | Y | Y | A | P | A | A | A | P | A | A | A | P | A | P | P | A | P | A | P | P | A | A | A | P | AP AA PA AA AA YY | AP PP PA AP AP AP HN | PA PP PP PA | 0 | IS04 | IS01 | SE |
| FID_Example4 | IID_Example4 | 2 | 1 | P | A | A | P | A | P | N | N | H | Y | A | P | A | A | A | P | A | A | A | P | A | P | A | A | P | A | A | P | A | A | A | P | AP AA AA AA AA HY | AP AP AA AP AP AP NN | PA AP AP PA | 0 | 0 | IS01 | SE |
| FID_Example5 | IID_Example5 | 2 | 1 | P | P | A | P | A | P | N | N | Y | Y | A | P | A | A | A | A | A | A | A | A | A | P | A | A | P | A | P | P | A | A | A | A | AA AA AA AA AA YY | AA PP AA AP AA AP NN | PA AP AP PP | 0 | 0 | IS01 | SE |
| | | | | | | | | | | | | | | | | | | | | | | | | | | | | | | | | | | | | | | | | | | N |
| | | | | | | | | | | | | | | | | | | | | | | | | | | | | | | | | | | | | | | | | | | N |
| | | | | | | | | | | | | | | | | | | | | | | | | | | | | | | | | | | | | | | | | | | N |
| | | | | | | | | | | | | | | | | | | | | | | | | | | | | | | | | | | | | | | | | | | N |
| | | | | | | | | | | | | | | | | | | | | | | | | | | | | | | | | | | | | | | | | | | N |
| | | | | | | | | | | | | | | | | | | | | | | | | | | | | | | | | | | | | | | | | | | N |
| | | | | | | | | | | | | | | | | | | | | | | | | | | | | | | | | | | | | | | | | | | N |
| | | | | | | | | | | | | | | | | | | | | | | | | | | | | | | | | | | | | | | | | | | N |
| | | | | | | | | | | | | | | | | | | | | | | | | | | | | | | | | | | | | | | | | | | N |
| | | | | | | | | | | | | | | | | | | | | | | | | | | | | | | | | | | | | | | | | | | N |
| | | | | | | | | | | | | | | | | | | | | | | | | | | | | | | | | | | | | | | | | | | N |
| | | | | | | | | | | | | | | | | | | | | | | | | | | | | | | | | | | | | | | | | | | N |
| | | | | | | | | | | | | | | | | | | | | | | | | | | | | | | | | | | | | | | | | | | N |
| | | | | | | | | | | | | | | | | | | | | | | | | | | | | | | | | | | | | | | | | | | N |
| | | | | | | | | | | | | | | | | | | | | | | | | | | | | | | | | | | | | | | | | | | N |
| | | | | | | | | | | | | | | | | | | | | | | | | | | | | | | | | | | | | | | | | | | N |
| | | | | | | | | | | | | | | | | | | | | | | | | | | | | | | | | | | | | | | | | | | N |
| | | | | | | | | | | | | | | | | | | | | | | | | | | | | | | | | | | | | | | | | | | N |
| | | | | | | | | | | | | | | | | | | | | | | | | | | | | | | | | | | | | | | | | | | N |
| | | | | | | | | | | | | | | | | | | | | | | | | | | | | | | | | | | | | | | | | | | N |
| | | | | | | | | | | | | | | | | | | | | | | | | | | | | | | | | | | | | | | | | | | N |
| | | | | | | | | | | | | | | | | | | | | | | | | | | | | | | | | | | | | | | | | | | N |
| | | | | | | | | | | | | | | | | | | | | | | | | | | | | | | | | | | | | | | | | | | N |
| | | | | | | | | | | | | | | | | | | | | | | | | | | | | | | | | | | | | | | | | | | N |
| | | | | | | | | | | | | | | | | | | | | | | | | | | | | | | | | | | | | | | | | | | N |
| | | | | | | | | | | | | | | | | | | | | | | | | | | | | | | | | | | | | | | | | | | N |
| | | | | | | | | | | | | | | | | | | | | | | | | | | | | | | | | | | | | | | | | | | N |
| | | | | | | | | | | | | | | | | | | | | | | | | | | | | | | | | | | | | | | | | | | N |
| | | | | | | | | | | | | | | | | | | | | | | | | | | | | | | | | | | | | | | | | | | N |
| | | | | | | | | | | | | | | | | | | | | | | | | | | | | | | | | | | | | | | | | | | N |
| | | | | | | | | | | | | | | | | | | | | | | | | | | | | | | | | | | | | | | | | | | N |
| | | | | | | | | | | | | | | | | | | | | | | | | | | | | | | | | | | | | | | | | | | N |
| | | | | | | | | | | | | | | | | | | | | | | | | | | | | | | | | | | | | | | | | | | N |

GuesSE is the algorythm for inferring HLA shared-epitope status from the output of the HLA imputation software SNP2HLA v1.0.3.
Usage:
* Use plink to extract the required  amino-acid calls from the SNP2HLA in .ped format.
*Paste the corresponding data in the GuesSE -- two columns for each imputed amino-acid call (one per "allele"). NB! The order of input AA data should match the order in the sheet. The input should begin with the standard .ped fields "IID","FID","GENDER" and "CASE" (see example entries).
* The output data is contained in columns "IF10", "IF04", "IF01" and "CALL" (where IF10, IF04, IF01 reflect imputed HLA types DRB1:10, DRB1:04 and DRB1:01; the "CALL" column represents the shared epitope positivity status of the individual).

**Required AA positions from SNP2HLA (extracted as a plink .ped file)**

AA_DRB1_71_32659935_R
AA_DRB1_70_32659938_Q
AA_DRB1_67_32659947_L
AA_DRB1_33_32660049
AA_DRB1_32_32660052
AA_DRB1_31_32660055_F
AA_DRB1_31_32660055_V
AA_DRB1_30_32660058_Y
AA_DRB1_30_32660058_R
AA_DRB1_28_32660064_D
AA_DRB1_26_32660070_F
AA_DRB1_11_32660115_V
AA_DRB1_11_32660115_L
AA_DRB1_10_32660118_Q
AA_DRB1_10_32660118_E
AA_DRB1_9_32660121_E
