## Supplementary Data Set 1 for "Identification of loci genetically associated with antibodies to specific sets of citrullinated peptides in rheumatoid arthritis patients"

| CHR | BP | SNP | A1 | A2 | N | P | P.R. | OR | OR.R. | Q | I | FDR | PEPTIDE |
|---|---|---|---|---|---|---|---|---|---|---|---|---|---|
| 1 | 113877958 | rs11102648 | A | C | 3 | 1.51E-05 | 0.0004994 | 1.1918 | 1.19 | 0.2207 | 33.82 | 0.098660569 | cfc1-cyc (CCP1) |
| 1 | 113926782 | rs1217203 | A | G | 2 | 1.48E-05 | 0.0001138 | 1.2413 | 1.2395 | 0.2663 | 19.09 | 0.098660569 | cfc1-cyc (CCP1) |
| 1 | 113953370 | rs17031716 | A | C | 3 | 4.19E-06 | 0.001685 | 1.2112 | 1.2092 | 0.1233 | 52.22 | 0.066780098 | cfc1-cyc (CCP1) |
| 1 | 113987113 | rs1230661 | A | G | 3 | 4.22E-06 | 0.0001062 | 1.2073 | 1.2062 | 0.2493 | 28.02 | 0.066780098 | cfc1-cyc (CCP1) |
| 1 | 114035394 | rs4839335 | G | A | 3 | 1.13E-05 | 0.0001215 | 1.1925 | 1.1912 | 0.2765 | 22.21 | 0.081865109 | cfc1-cyc (CCP1) |
| 1 | 114045700 | rs1230649 | C | A | 3 | 1.36E-05 | 0.0001919 | 1.1906 | 1.1891 | 0.2628 | 25.17 | 0.093049371 | cfc1-cyc (CCP1) |
| 1 | 114047976 | rs1936398 | G | A | 3 | 1.37E-05 | 0.0002426 | 1.1905 | 1.189 | 0.252 | 27.45 | 0.093105447 | cfc1-cyc (CCP1) |
| 1 | 114050905 | rs6698586 | G | A | 2 | 8.91E-06 | 7.34E-05 | 1.244 | 1.2423 | 0.2669 | 18.86 | 0.073315707 | cfc1-cyc (CCP1) |
| 1 | 114055162 | rs1230647 | G | A | 3 | 1.29E-05 | 0.0003345 | 1.1912 | 1.1894 | 0.2353 | 30.89 | 0.0898954 | cfc1-cyc (CCP1) |
| 1 | 114055415 | rs2295853 | A | G | 3 | 1.09E-05 | 0.0002679 | 1.193 | 1.1913 | 0.2398 | 29.98 | 0.081536019 | cfc1-cyc (CCP1) |
| 1 | 114068870 | rs1230640 | G | A | 3 | 1.53E-05 | 0.0006829 | 1.1894 | 1.1871 | 0.2064 | 36.63 | 0.098747513 | cfc1-cyc (CCP1) |
| 1 | 114069168 | rs1981319 | C | A | 3 | 1.30E-05 | 0.0002777 | 1.1911 | 1.1894 | 0.244 | 29.1 | 0.0898954 | cfc1-cyc (CCP1) |
| 1 | 114076960 | rs12061482 | A | G | 3 | 1.23E-05 | 0.000189 | 1.1917 | 1.1904 | 0.2595 | 25.87 | 0.087125999 | cfc1-cyc (CCP1) |
| 1 | 114078853 | rs12063216 | A | G | 3 | 1.09E-05 | 0.0002679 | 1.193 | 1.1913 | 0.2398 | 29.98 | 0.081536019 | cfc1-cyc (CCP1) |
| 1 | 114089425 | rs10858017 | G | A | 3 | 1.54E-05 | 0.0006176 | 1.1891 | 1.1871 | 0.2109 | 35.75 | 0.0988401 | cfc1-cyc (CCP1) |
| 1 | 114094067 | rs1230685 | G | A | 3 | 1.50E-05 | 0.0004948 | 1.1894 | 1.1876 | 0.2208 | 33.8 | 0.098660569 | cfc1-cyc (CCP1) |
| 1 | 114101039 | rs2359167 | A | G | 3 | 1.49E-06 | 0.000141 | 1.2181 | 1.2167 | 0.208 | 36.31 | 0.066780098 | cfc1-cyc (CCP1) |
| 1 | 114102858 | rs61817589 | A | G | 3 | 2.10E-06 | 0.0003349 | 1.2141 | 1.2125 | 0.1799 | 41.71 | 0.066780098 | cfc1-cyc (CCP1) |
| 1 | 114104489 | rs2797412 | A | G | 3 | 1.02E-05 | 0.0002981 | 1.1936 | 1.1919 | 0.2328 | 31.4 | 0.078792018 | cfc1-cyc (CCP1) |
| 1 | 114105331 | rs6679677 | A | C | 3 | 4.53E-07 | 4.53E-07 | 1.2842 | 1.2842 | 0.5692 | 0 | 0.049374952 | cfc1-cyc (CCP1) |
| 1 | 114113291 | rs10858018 | A | G | 3 | 1.18E-05 | 0.0001864 | 1.1922 | 1.1907 | 0.259 | 25.98 | 0.084712455 | cfc1-cyc (CCP1) |
| 1 | 114113958 | rs4145859 | G | C | 3 | 1.50E-06 | 0.0001104 | 1.2178 | 1.2164 | 0.2181 | 34.32 | 0.066780098 | cfc1-cyc (CCP1) |
| 1 | 114117743 | rs12041547 | G | C | 3 | 2.23E-06 | 0.0002053 | 1.2137 | 1.2122 | 0.203 | 37.3 | 0.066780098 | cfc1-cyc (CCP1) |
| 1 | 114139759 | rs1217396 | G | A | 3 | 1.51E-05 | 0.0002022 | 1.1896 | 1.1882 | 0.2635 | 25.01 | 0.098660569 | cfc1-cyc (CCP1) |
| 1 | 114159273 | rs1217413 | G | A | 3 | 2.62E-06 | 0.0002386 | 1.2122 | 1.2109 | 0.2003 | 37.8 | 0.066780098 | cfc1-cyc (CCP1) |
| 1 | 114167275 | rs1217389 | G | A | 3 | 1.51E-05 | 0.0002017 | 1.1896 | 1.1883 | 0.2637 | 24.98 | 0.098660569 | cfc1-cyc (CCP1) |
| 1 | 114175958 | rs1217395 | G | A | 3 | 9.46E-06 | 0.0001234 | 1.1942 | 1.1932 | 0.2688 | 23.88 | 0.075682517 | cfc1-cyc (CCP1) |
| 1 | 114178616 | rs1599971 | A | G | 3 | 1.57E-05 | 0.0002237 | 1.1892 | 1.1879 | 0.2601 | 25.73 | 0.099953135 | cfc1-cyc (CCP1) |
| 1 | 114179091 | rs2476601 | A | G | 3 | 4.02E-07 | 4.02E-07 | 1.2856 | 1.2856 | 0.5881 | 0 | 0.049374952 | cfc1-cyc (CCP1) |
| 1 | 114216891 | rs2488457 | G | C | 3 | 5.14E-06 | 0.0007875 | 1.208 | 1.2074 | 0.1615 | 45.15 | 0.066780098 | cfc1-cyc (CCP1) |
| 1 | 114221851 | rs12566340 | A | G | 3 | 1.15E-05 | 0.0001874 | 1.1954 | 1.1946 | 0.2555 | 26.71 | 0.083354424 | cfc1-cyc (CCP1) |
| 1 | 114221985 | rs7529353 | A | G | 3 | 9.66E-06 | 0.0001955 | 1.1971 | 1.1962 | 0.2481 | 28.26 | 0.076214959 | cfc1-cyc (CCP1) |
| 1 | 114230984 | rs2358994 | A | G | 3 | 5.79E-06 | 5.79E-06 | 1.2213 | 1.2213 | 0.6524 | 0 | 0.070518994 | cfc1-cyc (CCP1) |
| 1 | 114231038 | rs2358995 | C | A | 3 | 6.33E-06 | 6.33E-06 | 1.2202 | 1.2202 | 0.664 | 0 | 0.070518994 | cfc1-cyc (CCP1) |
| 1 | 114249912 | rs11552449 | A | G | 3 | 3.92E-06 | 3.92E-06 | 1.2275 | 1.2275 | 0.5553 | 0 | 0.066780098 | cfc1-cyc (CCP1) |
| 1 | 114251185 | rs3761936 | G | A | 3 | 4.05E-06 | 4.05E-06 | 1.2268 | 1.2268 | 0.5459 | 0 | 0.066780098 | cfc1-cyc (CCP1) |
| 1 | 114251352 | rs11102701 | T | A | 3 | 2.29E-06 | 2.29E-06 | 1.2345 | 1.2345 | 0.5568 | 0 | 0.066780098 | cfc1-cyc (CCP1) |
| 11 | 23421508 | rs7113623 | A | G | 2 | 1.22E-05 | 1.22E-05 | 1.3379 | 1.3379 | 0.8974 | 0 | 0.08675539 | cfc1-cyc (CCP1) |
| 11 | 23421835 | rs4528328 | C | A | 2 | 1.22E-05 | 1.22E-05 | 1.3379 | 1.3379 | 0.8974 | 0 | 0.08675539 | cfc1-cyc (CCP1) |
| 11 | 23422881 | rs1564102 | G | A | 3 | 4.96E-06 | 4.96E-06 | 1.2747 | 1.2747 | 0.4676 | 0 | 0.066780098 | cfc1-cyc (CCP1) |
| 11 | 23423031 | rs1564100 | G | A | 3 | 8.47E-06 | 8.47E-06 | 1.2673 | 1.2673 | 0.4793 | 0 | 0.071744742 | cfc1-cyc (CCP1) |
| 11 | 23426531 | rs2126507 | G | A | 3 | 5.20E-06 | 5.20E-06 | 1.2742 | 1.2742 | 0.4717 | 0 | 0.066780098 | cfc1-cyc (CCP1) |
| 1 | 113926782 | rs1217203 | A | G | 2 | 3.47E-06 | 3.47E-06 | 1.2609 | 1.2609 | 0.6878 | 0 | 0.066780098 | CEP-1 |
| 1 | 113962281 | rs11102661 | A | G | 3 | 6.79E-06 | 0.003546 | 1.2051 | 1.1996 | 0.1063 | 55.38 | 0.070518994 | CEP-1 |
| 1 | 113962639 | rs1217200 | C | A | 3 | 7.78E-06 | 0.00587 | 1.2033 | 1.1975 | 0.0847 | 59.49 | 0.070518994 | CEP-1 |
| 1 | 113964142 | rs1217193 | C | G | 3 | 7.62E-06 | 0.006655 | 1.2037 | 1.1974 | 0.079 | 60.61 | 0.070518994 | CEP-1 |
| 1 | 113966031 | rs12565589 | A | G | 3 | 8.82E-06 | 0.006522 | 1.202 | 1.1961 | 0.0823 | 59.96 | 0.073315707 | CEP-1 |
| 1 | 113972360 | rs1777234 | A | T | 3 | 7.67E-06 | 0.006708 | 1.2036 | 1.1974 | 0.0786 | 60.68 | 0.070518994 | CEP-1 |
| 1 | 113987113 | rs1230661 | A | G | 3 | 4.97E-06 | 0.02649 | 1.2126 | 1.2032 | 0.0214 | 73.99 | 0.066780098 | CEP-1 |
| 1 | 113988894 | rs1777237 | G | A | 3 | 9.38E-06 | 0.005715 | 1.2013 | 1.1956 | 0.0897 | 58.53 | 0.075581721 | CEP-1 |
| 1 | 113992696 | rs1777238 | G | A | 3 | 9.38E-06 | 0.005715 | 1.2013 | 1.1956 | 0.0897 | 58.53 | 0.075581721 | CEP-1 |
| 1 | 113998406 | rs1146182 | C | A | 3 | 9.20E-06 | 0.005745 | 1.2015 | 1.1959 | 0.089 | 58.67 | 0.074908787 | CEP-1 |
| 1 | 114008837 | rs1743605 | C | A | 3 | 8.35E-06 | 0.004853 | 1.2025 | 1.1969 | 0.0954 | 57.44 | 0.071744742 | CEP-1 |
| 1 | 114014094 | rs1146187 | A | G | 3 | 8.62E-06 | 0.005816 | 1.2023 | 1.1965 | 0.0872 | 59.02 | 0.072538889 | CEP-1 |
| 1 | 114019784 | rs1230659 | A | G | 3 | 7.20E-06 | 0.003556 | 1.2042 | 1.1991 | 0.1068 | 55.29 | 0.070518994 | CEP-1 |
| 1 | 114019974 | rs1230658 | C | G | 3 | 7.81E-06 | 0.003495 | 1.2033 | 1.1984 | 0.1092 | 54.84 | 0.070518994 | CEP-1 |
| 1 | 114035394 | rs4839335 | G | A | 3 | 6.53E-06 | 0.003656 | 1.2051 | 1.1994 | 0.1044 | 55.75 | 0.070518994 | CEP-1 |
| 1 | 114045700 | rs1230649 | C | A | 3 | 5.70E-06 | 0.003479 | 1.2066 | 1.201 | 0.1036 | 55.89 | 0.070327765 | CEP-1 |
| 1 | 114047976 | rs1936398 | G | A | 3 | 5.25E-06 | 0.003558 | 1.2074 | 1.2017 | 0.101 | 56.37 | 0.066780098 | CEP-1 |
| 1 | 114050905 | rs6698586 | G | A | 2 | 3.76E-07 | 3.76E-07 | 1.2841 | 1.2841 | 0.9438 | 0 | 0.049374952 | CEP-1 |
| 1 | 114055162 | rs1230647 | G | A | 3 | 4.89E-06 | 0.004946 | 1.2083 | 1.2022 | 0.0842 | 59.59 | 0.066780098 | CEP-1 |
| 1 | 114055415 | rs2295853 | A | G | 3 | 4.99E-06 | 0.004922 | 1.208 | 1.2019 | 0.0848 | 59.46 | 0.066780098 | CEP-1 |
| 1 | 114068870 | rs1230640 | G | A | 3 | 4.91E-06 | 0.00444 | 1.2082 | 1.202 | 0.0897 | 58.53 | 0.066780098 | CEP-1 |
| 1 | 114069168 | rs1981319 | C | A | 3 | 5.95E-06 | 0.004791 | 1.2061 | 1.2002 | 0.0895 | 58.56 | 0.070518994 | CEP-1 |
| 1 | 114076960 | rs12061482 | A | G | 3 | 4.87E-06 | 0.00382 | 1.2083 | 1.2028 | 0.0955 | 57.43 | 0.066780098 | CEP-1 |
| 1 | 114078853 | rs12063216 | A | G | 3 | 6.02E-06 | 0.004827 | 1.206 | 1.1999 | 0.0896 | 58.54 | 0.070518994 | CEP-1 |
| 1 | 114089425 | rs10858017 | G | A | 3 | 5.23E-06 | 0.004303 | 1.2073 | 1.2013 | 0.092 | 58.09 | 0.066780098 | CEP-1 |
| 1 | 114094067 | rs1230685 | G | A | 3 | 4.87E-06 | 0.004678 | 1.2081 | 1.2019 | 0.087 | 59.05 | 0.066780098 | CEP-1 |
| 1 | 114101039 | rs2359167 | A | G | 3 | 2.55E-06 | 0.02188 | 1.2201 | 1.2099 | 0.0222 | 73.73 | 0.066780098 | CEP-1 |
| 1 | 114102858 | rs61817589 | A | G | 3 | 2.74E-06 | 0.02428 | 1.2187 | 1.2087 | 0.0197 | 74.55 | 0.066780098 | CEP-1 |
| 1 | 114104489 | rs2797412 | A | G | 3 | 6.72E-06 | 0.004719 | 1.2049 | 1.199 | 0.0927 | 57.96 | 0.070518994 | CEP-1 |
| 1 | 114105331 | rs6679677 | A | C | 3 | 7.87E-09 | 9.00E-09 | 1.3469 | 1.3468 | 0.3653 | 0.7 | 0.002963304 | CEP-1 |
| 1 | 114113291 | rs10858018 | A | G | 3 | 5.29E-06 | 0.003743 | 1.2074 | 1.2018 | 0.0986 | 56.84 | 0.066780098 | CEP-1 |
| 1 | 114113958 | rs4145859 | G | C | 3 | 2.91E-06 | 0.02121 | 1.2184 | 1.2086 | 0.0238 | 73.26 | 0.066780098 | CEP-1 |
| 1 | 114117743 | rs12041547 | G | C | 3 | 3.91E-06 | 0.02347 | 1.2151 | 1.2053 | 0.0233 | 73.4 | 0.066780098 | CEP-1 |
| 1 | 114139759 | rs1217396 | G | A | 3 | 7.19E-06 | 0.003555 | 1.2042 | 1.1991 | 0.1068 | 55.29 | 0.070518994 | CEP-1 |
| 1 | 114144859 | rs1217380 | G | A | 3 | 7.65E-06 | 0.004025 | 1.2036 | 1.1983 | 0.1022 | 56.15 | 0.070518994 | CEP-1 |
| 1 | 114158734 | rs1217412 | G | A | 3 | 8.18E-06 | 0.004542 | 1.2029 | 1.1975 | 0.0978 | 56.98 | 0.071180056 | CEP-1 |
| 1 | 114159273 | rs1217413 | G | A | 3 | 4.54E-06 | 0.02148 | 1.2137 | 1.2047 | 0.0266 | 72.43 | 0.066780098 | CEP-1 |
| 1 | 114165999 | rs1217388 | G | A | 3 | 7.65E-06 | 0.004025 | 1.2036 | 1.1983 | 0.1022 | 56.15 | 0.070518994 | CEP-1 |
| 1 | 114167275 | rs1217389 | G | A | 3 | 6.73E-06 | 0.00311 | 1.205 | 1.2 | 0.1119 | 54.34 | 0.070518994 | CEP-1 |
| 1 | 114175958 | rs1217395 | G | A | 3 | 7.06E-06 | 0.005044 | 1.2042 | 1.1991 | 0.0886 | 58.73 | 0.070518994 | CEP-1 |
| 1 | 114178616 | rs1599971 | A | G | 3 | 6.73E-06 | 0.003108 | 1.205 | 1.2 | 0.1119 | 54.34 | 0.070518994 | CEP-1 |
| 1 | 114179091 | rs2476601 | A | G | 3 | 5.40E-09 | 5.40E-09 | 1.3514 | 1.3514 | 0.4203 | 0 | 0.002709866 | CEP-1 |
| 1 | 114195271 | rs1217407 | A | G | 3 | 7.68E-06 | 0.00461 | 1.2036 | 1.1982 | 0.0958 | 57.36 | 0.070518994 | CEP-1 |
| 1 | 114207974 | rs2488458 | A | G | 3 | 9.84E-06 | 0.006062 | 1.2008 | 1.195 | 0.0881 | 58.84 | 0.076798804 | CEP-1 |
| 1 | 114216891 | rs2488457 | G | C | 3 | 4.87E-06 | 0.01218 | 1.2158 | 1.2101 | 0.0442 | 67.94 | 0.066780098 | CEP-1 |
| 1 | 114221851 | rs12566340 | A | G | 3 | 2.43E-06 | 0.0007449 | 1.2191 | 1.2163 | 0.1514 | 47.02 | 0.066780098 | CEP-1 |
| 1 | 114221985 | rs7529353 | A | G | 3 | 2.30E-06 | 0.0008578 | 1.2196 | 1.2167 | 0.1436 | 48.47 | 0.066780098 | CEP-1 |
| 1 | 114222836 | rs11102691 | A | G | 3 | 4.98E-06 | 0.0009047 | 1.2132 | 1.2104 | 0.1609 | 45.27 | 0.066780098 | CEP-1 |
| 1 | 114230984 | rs2358994 | A | G | 3 | 6.57E-06 | 6.57E-06 | 1.2286 | 1.2286 | 0.7327 | 0 | 0.070518994 | CEP-1 |
| 1 | 114231038 | rs2358995 | C | A | 3 | 7.23E-06 | 7.23E-06 | 1.2274 | 1.2274 | 0.7386 | 0 | 0.070518994 | CEP-1 |
| 1 | 114249912 | rs11552449 | A | G | 3 | 7.59E-06 | 7.59E-06 | 1.2288 | 1.2288 | 0.7653 | 0 | 0.070518994 | CEP-1 |
| 1 | 114251185 | rs3761936 | G | A | 3 | 8.10E-06 | 8.10E-06 | 1.2277 | 1.2277 | 0.7733 | 0 | 0.071180056 | CEP-1 |
| 1 | 114251352 | rs11102701 | T | A | 3 | 4.32E-06 | 4.32E-06 | 1.2367 | 1.2367 | 0.7621 | 0 | 0.066780098 | CEP-1 |
| 13 | 41850075 | rs12871509 | C | A | 3 | 3.96E-06 | 3.96E-06 | 1.2377 | 1.2377 | 0.5463 | 0 | 0.066780098 | CEP-1 |
| 13 | 41850145 | rs9594738 | A | G | 3 | 1.30E-05 | 0.0004802 | 0.8495 | 0.8496 | 0.2128 | 35.37 | 0.0898954 | CEP-1 |
| 13 | 41850499 | rs17457484 | A | G | 3 | 3.61E-06 | 3.61E-06 | 1.2389 | 1.2389 | 0.4921 | 0 | 0.066780098 | CEP-1 |
| 13 | 41850611 | rs56008941 | A | G | 3 | 4.58E-06 | 4.58E-06 | 1.2361 | 1.2361 | 0.513 | 0 | 0.066780098 | CEP-1 |
| 13 | 41851003 | rs1475248 | A | G | 3 | 4.23E-06 | 4.23E-06 | 1.2369 | 1.2369 | 0.5442 | 0 | 0.066780098 | CEP-1 |
| 13 | 41851336 | rs989354 | G | A | 3 | 4.81E-06 | 4.81E-06 | 1.2356 | 1.2356 | 0.5471 | 0 | 0.066780098 | CEP-1 |
| 13 | 41853965 | rs12870516 | C | G | 3 | 3.99E-06 | 3.99E-06 | 1.2379 | 1.2379 | 0.548 | 0 | 0.066780098 | CEP-1 |
| 13 | 41854220 | rs35726267 | G | A | 3 | 4.40E-06 | 4.40E-06 | 1.2366 | 1.2366 | 0.5491 | 0 | 0.066780098 | CEP-1 |
| 13 | 41854242 | rs35469241 | G | A | 3 | 4.40E-06 | 4.40E-06 | 1.2366 | 1.2366 | 0.5491 | 0 | 0.066780098 | CEP-1 |
| 13 | 41855324 | rs35406824 | A | G | 3 | 5.32E-06 | 5.32E-06 | 1.2342 | 1.2342 | 0.5649 | 0 | 0.066780098 | CEP-1 |
| 13 | 41855732 | rs61960917 | G | A | 3 | 6.14E-06 | 6.14E-06 | 1.2324 | 1.2324 | 0.5481 | 0 | 0.070518994 | CEP-1 |
| 13 | 41856193 | rs56737241 | A | C | 3 | 5.32E-06 | 5.32E-06 | 1.2342 | 1.2342 | 0.5649 | 0 | 0.066780098 | CEP-1 |
| 13 | 41867646 | rs10507507 | G | A | 3 | 6.58E-06 | 6.58E-06 | 1.2323 | 1.2323 | 0.5454 | 0 | 0.070518994 | CEP-1 |
| 13 | 41868202 | rs17535675 | C | A | 3 | 6.87E-06 | 6.87E-06 | 1.2311 | 1.2311 | 0.536 | 0 | 0.070518994 | CEP-1 |
| 13 | 41869543 | rs12855879 | A | C | 3 | 8.39E-06 | 8.39E-06 | 1.2288 | 1.2288 | 0.5589 | 0 | 0.071744742 | CEP-1 |
| 13 | 41872818 | rs9533096 | A | G | 3 | 3.51E-06 | 3.51E-06 | 1.2383 | 1.2383 | 0.4928 | 0 | 0.066780098 | CEP-1 |
| 13 | 41875809 | rs12874142 | A | G | 3 | 1.12E-05 | 1.12E-05 | 1.2257 | 1.2257 | 0.5752 | 0 | 0.081865109 | CEP-1 |
| 13 | 41876035 | rs12853439 | A | G | 3 | 9.73E-06 | 9.73E-06 | 1.2275 | 1.2275 | 0.5981 | 0 | 0.076351494 | CEP-1 |
| 13 | 41877444 | rs12865514 | G | A | 3 | 7.33E-06 | 7.33E-06 | 1.2256 | 1.2256 | 0.6574 | 0 | 0.070518994 | CEP-1 |
| 13 | 41886924 | rs7327510 | G | A | 3 | 3.78E-06 | 0.0003521 | 1.1906 | 1.1892 | 0.1939 | 39.05 | 0.066780098 | CEP-1 |
| 13 | 41890135 | rs9525619 | G | A | 3 | 8.02E-06 | 0.0001102 | 1.1835 | 1.1829 | 0.2676 | 24.14 | 0.071180056 | CEP-1 |
| 13 | 41890977 | rs9315913 | G | A | 3 | 4.88E-06 | 4.88E-06 | 1.2316 | 1.2316 | 0.6684 | 0 | 0.066780098 | CEP-1 |
| 13 | 41891791 | rs9315914 | A | G | 3 | 3.57E-06 | 3.57E-06 | 1.2354 | 1.2354 | 0.6885 | 0 | 0.066780098 | CEP-1 |

| | | | | | | | | | | | | | |
|---|---|---|---|---|---|---|---|---|---|---|---|---|---|
| 13 | 41892481 | rs9590697 | G | C | 3 | 6.80E-06 | 6.80E-06 | 1.2289 | 1.2289 | 0.6745 | 0 | 0.070518994 | CEP-1 |
| 13 | 41893262 | rs727243 | A | G | 3 | 4.13E-06 | 4.13E-06 | 1.2336 | 1.2336 | 0.6772 | 0 | 0.066780098 | CEP-1 |
| 13 | 41895324 | rs912425 | G | A | 3 | 2.20E-06 | 2.20E-06 | 1.2376 | 1.2376 | 0.8985 | 0 | 0.066780098 | CEP-1 |
| 13 | 41903533 | rs4942120 | G | A | 3 | 3.98E-06 | 3.98E-06 | 1.2265 | 1.2265 | 0.931 | 0 | 0.066780098 | CEP-1 |
| 13 | 41905194 | rs2324873 | G | A | 3 | 3.30E-06 | 3.30E-06 | 1.2311 | 1.2311 | 0.8532 | 0 | 0.066780098 | CEP-1 |
| 13 | 41906177 | rs4941428 | A | G | 3 | 2.80E-06 | 2.80E-06 | 1.2306 | 1.2306 | 0.9219 | 0 | 0.066780098 | CEP-1 |
| 13 | 41906994 | rs1924416 | G | A | 3 | 4.21E-06 | 4.21E-06 | 1.2288 | 1.2288 | 0.9123 | 0 | 0.066780098 | CEP-1 |
| 13 | 41907008 | rs1924415 | C | G | 3 | 2.04E-06 | 2.04E-06 | 1.2416 | 1.2416 | 0.8286 | 0 | 0.066780098 | CEP-1 |
| 13 | 41907566 | rs1575525 | G | C | 3 | 4.38E-06 | 4.38E-06 | 1.2285 | 1.2285 | 0.9364 | 0 | 0.066780098 | CEP-1 |
| 13 | 41909160 | rs9525624 | A | C | 3 | 1.02E-06 | 1.02E-06 | 1.2452 | 1.2452 | 0.9257 | 0 | 0.066780098 | CEP-1 |
| 13 | 41910590 | rs4942121 | A | G | 3 | 1.25E-06 | 1.25E-06 | 1.2462 | 1.2462 | 0.9102 | 0 | 0.066780098 | CEP-1 |
| 13 | 41912464 | rs12864265 | A | G | 3 | 2.25E-06 | 2.25E-06 | 1.2405 | 1.2405 | 0.8968 | 0 | 0.066780098 | CEP-1 |
| 13 | 41914294 | rs17535911 | C | A | 3 | 2.35E-06 | 2.35E-06 | 1.2399 | 1.2399 | 0.9099 | 0 | 0.066780098 | CEP-1 |
| 13 | 41915369 | rs6561045 | G | A | 3 | 1.47E-06 | 1.47E-06 | 1.2443 | 1.2443 | 0.8977 | 0 | 0.066780098 | CEP-1 |
| 13 | 41915470 | rs7316953 | A | G | 3 | 1.67E-06 | 1.67E-06 | 1.2429 | 1.2429 | 0.9117 | 0 | 0.066780098 | CEP-1 |
| 13 | 41915500 | rs1324005 | C | G | 3 | 2.38E-06 | 2.38E-06 | 1.2398 | 1.2398 | 0.8899 | 0 | 0.066780098 | CEP-1 |
| 13 | 41915895 | rs9533104 | C | G | 3 | 2.56E-06 | 2.56E-06 | 1.1939 | 1.1939 | 0.4092 | 0 | 0.066780098 | CEP-1 |
| 13 | 41916030 | rs9525625 | A | G | 3 | 2.20E-06 | 2.20E-06 | 1.1954 | 1.1954 | 0.4039 | 0 | 0.066780098 | CEP-1 |
| 13 | 41916177 | rs720824 | A | T | 3 | 2.05E-06 | 2.05E-06 | 1.2416 | 1.2416 | 0.8944 | 0 | 0.066780098 | CEP-1 |
| 13 | 41916898 | rs4942122 | G | A | 3 | 2.57E-06 | 2.57E-06 | 1.2389 | 1.2389 | 0.901 | 0 | 0.066780098 | CEP-1 |
| 13 | 41917046 | rs4942123 | G | A | 3 | 2.05E-06 | 2.05E-06 | 1.2416 | 1.2416 | 0.8944 | 0 | 0.066780098 | CEP-1 |
| 13 | 41932968 | rs1853573 | G | C | 3 | 1.03E-06 | 1.33E-05 | 1.2029 | 1.2028 | 0.2858 | 20.15 | 0.066780098 | CEP-1 |
| 13 | 41933209 | rs17536002 | G | A | 3 | 2.27E-06 | 2.27E-06 | 1.2404 | 1.2404 | 0.889 | 0 | 0.066780098 | CEP-1 |
| 13 | 41938043 | rs9594766 | G | A | 3 | 2.06E-06 | 2.07E-06 | 1.196 | 1.196 | 0.3678 | 0.02 | 0.066780098 | CEP-1 |
| 13 | 41944036 | rs2093816 | G | A | 3 | 3.63E-06 | 1.65E-05 | 1.1912 | 1.1911 | 0.3162 | 13.16 | 0.066780098 | CEP-1 |
| 13 | 41944054 | rs927623 | G | A | 3 | 2.62E-06 | 2.62E-06 | 1.2388 | 1.2388 | 0.8961 | 0 | 0.066780098 | CEP-1 |
| 13 | 41944812 | rs9533117 | A | T | 3 | 3.22E-06 | 3.22E-06 | 1.233 | 1.233 | 0.8534 | 0 | 0.066780098 | CEP-1 |
| 13 | 41945920 | rs9315920 | G | A | 3 | 3.72E-06 | 3.72E-06 | 1.2301 | 1.2301 | 0.9555 | 0 | 0.066780098 | CEP-1 |
| 13 | 41947482 | rs9594768 | C | A | 2 | 1.99E-06 | 1.99E-06 | 1.2439 | 1.2439 | 0.7297 | 0 | 0.066780098 | CEP-1 |
| 13 | 41949804 | rs912423 | T | A | 3 | 3.99E-06 | 3.99E-06 | 1.2293 | 1.2293 | 0.9604 | 0 | 0.066780098 | CEP-1 |
| 13 | 41950880 | rs2062305 | G | A | 3 | 2.10E-06 | 2.10E-06 | 1.196 | 1.196 | 0.3679 | 0 | 0.066780098 | CEP-1 |
| 13 | 41955549 | rs1351832 | A | G | 3 | 3.03E-06 | 3.03E-06 | 1.2297 | 1.2297 | 0.9569 | 0 | 0.066780098 | CEP-1 |
| 13 | 41958407 | rs17536071 | A | G | 3 | 1.88E-06 | 1.88E-06 | 1.2417 | 1.2417 | 0.8671 | 0 | 0.066780098 | CEP-1 |
| 13 | 41959963 | rs12871228 | A | G | 3 | 8.17E-06 | 8.17E-06 | 1.222 | 1.222 | 0.9512 | 0 | 0.071180056 | CEP-1 |
| 13 | 41962910 | rs12868231 | A | G | 3 | 8.89E-06 | 8.89E-06 | 1.2211 | 1.2211 | 0.9299 | 0 | 0.073315707 | CEP-1 |
| 1 | 7927550 | rs9434948 | C | A | 2 | 2.49E-06 | 2.49E-06 | 0.8143 | 0.8143 | 0.3945 | 0 | 0.066780098 | Cit-FibÎ²36-52 |
| 1 | 113987113 | rs1230661 | A | G | 3 | 1.06E-05 | 0.0001392 | 1.1973 | 1.1959 | 0.2686 | 23.93 | 0.080236034 | Cit-FibÎ²36-52 |
| 1 | 114101039 | rs2359167 | A | G | 3 | 6.45E-06 | 0.0001632 | 1.2027 | 1.2008 | 0.2462 | 28.65 | 0.070518994 | Cit-FibÎ²36-52 |
| 1 | 114102858 | rs61817589 | A | G | 3 | 1.12E-05 | 0.0004737 | 1.1964 | 1.1942 | 0.2144 | 35.05 | 0.081865109 | Cit-FibÎ²36-52 |
| 1 | 114105331 | rs6679677 | A | C | 3 | 1.82E-07 | 5.45E-05 | 1.2947 | 1.291 | 0.1975 | 38.35 | 0.034249906 | Cit-FibÎ²36-52 |
| 1 | 114113958 | rs4145859 | G | C | 3 | 7.22E-06 | 0.0001326 | 1.2013 | 1.1996 | 0.2586 | 26.06 | 0.070518994 | Cit-FibÎ²36-52 |
| 1 | 114117743 | rs12041547 | G | C | 3 | 1.06E-05 | 0.0002743 | 1.1972 | 1.1953 | 0.2386 | 30.22 | 0.080236034 | Cit-FibÎ²36-52 |
| 1 | 114159273 | rs1217413 | G | A | 3 | 9.62E-06 | 0.0001653 | 1.1983 | 1.1967 | 0.258 | 26.19 | 0.076214959 | Cit-FibÎ²36-52 |
| 1 | 114179091 | rs2476601 | A | G | 3 | 1.26E-07 | 1.70E-05 | 1.2991 | 1.296 | 0.2294 | 32.08 | 0.027049168 | Cit-FibÎ²36-52 |
| 1 | 114216891 | rs2488457 | G | C | 3 | 3.20E-06 | 0.000361 | 1.2124 | 1.2105 | 0.1888 | 40.01 | 0.066780098 | Cit-FibÎ²36-52 |
| 1 | 114221985 | rs7529353 | A | G | 3 | 1.53E-05 | 0.0007566 | 1.1918 | 1.1897 | 0.2002 | 37.82 | 0.098747513 | Cit-FibÎ²36-52 |
| 1 | 114105331 | rs6679677 | A | C | 3 | 2.98E-06 | 0.0001008 | 1.259 | 1.2557 | 0.2461 | 28.68 | 0.066780098 | Cit-FibÎ±563-583 |
| 1 | 114179091 | rs2476601 | A | G | 3 | 2.66E-06 | 7.53E-05 | 1.2604 | 1.2573 | 0.2544 | 26.95 | 0.066780098 | Cit-FibÎ±563-583 |
| 1 | 114105331 | rs6679677 | A | C | 3 | 1.37E-06 | 0.00246 | 1.2861 | 1.2872 | 0.0776 | 60.88 | 0.066780098 | Cit-FibÎ±580-600 |
| 1 | 114179091 | rs2476601 | A | G | 3 | 1.49E-06 | 0.002827 | 1.2849 | 1.286 | 0.0735 | 61.69 | 0.066780098 | Cit-FibÎ±580-600 |
| 1 | 114230984 | rs2358994 | A | G | 3 | 7.40E-07 | 2.41E-05 | 1.2581 | 1.2582 | 0.2527 | 27.31 | 0.063590404 | Cit-FibÎ±580-600 |
| 1 | 114231038 | rs2358995 | C | A | 3 | 8.02E-07 | 2.13E-05 | 1.2571 | 1.2573 | 0.2594 | 25.88 | 0.063590404 | Cit-FibÎ±580-600 |
| 1 | 114249912 | rs11552449 | A | G | 3 | 9.47E-07 | 8.93E-05 | 1.2573 | 1.2575 | 0.2086 | 36.19 | 0.066780098 | Cit-FibÎ±580-600 |
| 1 | 114251185 | rs3761936 | G | A | 3 | 6.93E-07 | 0.0001061 | 1.2606 | 1.2608 | 0.1939 | 39.05 | 0.063590404 | Cit-FibÎ±580-600 |
| 1 | 114251352 | rs11102701 | T | A | 3 | 4.59E-07 | 0.0001287 | 1.2666 | 1.267 | 0.1756 | 42.51 | 0.049374952 | Cit-FibÎ±580-600 |
| 11 | 6244494 | rs11040832 | G | A | 3 | 1.53E-05 | 1.53E-05 | 1.2272 | 1.2272 | 0.5472 | 0 | 0.098747513 | Cit-FibÎ±580-600 |
| 11 | 70896820 | rs117978959 | A | G | 2 | 1.05E-06 | 1.05E-06 | 1.628 | 1.628 | 0.4192 | 0 | 0.066780098 | Cit-FibÎ±36-50 |
| 17 | 12107064 | rs12452363 | A | G | 3 | 3.85E-06 | 3.85E-06 | 1.2608 | 1.2608 | 0.4358 | 0 | 0.066780098 | Cit-FibÎ±36-50 |
| 1 | 114105331 | rs6679677 | A | C | 3 | 7.37E-08 | 7.37E-08 | 1.3145 | 1.3145 | 0.7345 | 0 | 0.018495075 | Cit-FibÎ±621-635 |
| 1 | 114179091 | rs2476601 | A | G | 3 | 4.93E-08 | 4.93E-08 | 1.3193 | 1.3193 | 0.7282 | 0 | 0.014843258 | Cit-FibÎ±621-635 |
| 1 | 114216891 | rs2488457 | G | C | 3 | 1.04E-05 | 1.04E-05 | 1.2078 | 1.2078 | 0.9877 | 0 | 0.079619666 | Cit-FibÎ±621-635 |
| 1 | 114230984 | rs2358994 | A | G | 3 | 5.94E-06 | 5.94E-06 | 1.229 | 1.229 | 0.3732 | 0 | 0.070518994 | Cit-FibÎ±621-635 |
| 1 | 114231038 | rs2358995 | C | A | 3 | 6.48E-06 | 6.48E-06 | 1.228 | 1.228 | 0.3812 | 0 | 0.070518994 | Cit-FibÎ±621-635 |
| 1 | 114251185 | rs3761936 | G | A | 3 | 1.32E-05 | 0.0001577 | 1.2211 | 1.2182 | 0.2753 | 22.47 | 0.090447862 | Cit-FibÎ±621-635 |
| 1 | 114251352 | rs11102701 | T | A | 3 | 7.09E-06 | 1.85E-05 | 1.2296 | 1.2287 | 0.3367 | 8.14 | 0.070518994 | Cit-FibÎ±621-635 |
| 2 | 43386028 | rs41382648 | A | G | 3 | 7.58E-06 | 0.0001424 | 1.2935 | 1.2912 | 0.2561 | 26.58 | 0.070518994 | Cit-FibÎ±621-635 |
| 2 | 43419650 | rs4372955 | G | A | 3 | 8.48E-06 | 0.0004946 | 1.2898 | 1.2869 | 0.2021 | 37.46 | 0.071744742 | Cit-FibÎ±621-635 |
| 1 | 114101039 | rs2359167 | A | G | 3 | 8.12E-06 | 8.12E-06 | 1.2126 | 1.2126 | 0.6045 | 0 | 0.071180056 | Cit-FibÎ²60-74 |
| 1 | 114102858 | rs61817589 | A | G | 3 | 1.29E-05 | 1.29E-05 | 1.2069 | 1.2069 | 0.5855 | 0 | 0.0898954 | Cit-FibÎ²60-74 |
| 1 | 114105331 | rs6679677 | A | C | 3 | 7.83E-07 | 3.21E-06 | 1.2968 | 1.2964 | 0.3272 | 10.49 | 0.063590404 | Cit-FibÎ²60-74 |
| 1 | 114113958 | rs4145859 | G | C | 3 | 7.87E-06 | 7.87E-06 | 1.2128 | 1.2128 | 0.6029 | 0 | 0.070518994 | Cit-FibÎ²60-74 |
| 1 | 114117743 | rs12041547 | G | C | 3 | 7.62E-06 | 7.62E-06 | 1.2131 | 1.2131 | 0.5707 | 0 | 0.070518994 | Cit-FibÎ²60-74 |
| 1 | 114159273 | rs1217413 | G | A | 3 | 9.12E-06 | 9.12E-06 | 1.2112 | 1.2112 | 0.6198 | 0 | 0.074628233 | Cit-FibÎ²60-74 |
| 1 | 114179091 | rs2476601 | A | G | 3 | 4.94E-07 | 4.94E-07 | 1.3026 | 1.3026 | 0.392 | 0 | 0.049648241 | Cit-FibÎ²60-74 |
| 1 | 114216891 | rs2488457 | G | C | 3 | 6.67E-06 | 6.67E-06 | 1.2177 | 1.2177 | 0.4894 | 0 | 0.070518994 | Cit-FibÎ²60-74 |
| 17 | 13383091 | rs12941565 | A | G | 3 | 6.77E-06 | 6.77E-06 | 0.805 | 0.805 | 0.5997 | 0 | 0.070518994 | Cit-FibÎ²60-74 |
| 2 | 181864898 | rs61358146 | A | G | 2 | 1.41E-05 | 1.41E-05 | 0.7916 | 0.7916 | 0.9038 | 0 | 0.095445172 | Cit-C1(Cit-R) |
| 2 | 181865035 | rs6728366 | A | C | 2 | 1.49E-05 | 1.49E-05 | 0.792 | 0.792 | 0.8914 | 0 | 0.098660569 | Cit-C1(Cit-R) |
| 2 | 181869724 | rs1899036 | A | T | 2 | 1.29E-05 | 1.29E-05 | 0.7904 | 0.7904 | 0.7159 | 0 | 0.0898954 | Cit-C1(Cit-R) |
| 5 | 72509655 | rs6893265 | G | A | 3 | 5.14E-06 | 5.14E-06 | 0.7288 | 0.7288 | 0.5526 | 0 | 0.066780098 | Cit-C1(Cit-R) |
| 5 | 72514645 | rs73762301 | G | A | 3 | 6.79E-06 | 6.79E-06 | 0.7321 | 0.7321 | 0.5384 | 0 | 0.070518994 | Cit-C1(Cit-R) |
| 5 | 72515584 | rs73762302 | G | C | 3 | 7.32E-06 | 7.32E-06 | 0.7329 | 0.7329 | 0.5388 | 0 | 0.070518994 | Cit-C1(Cit-R) |
| 5 | 72517327 | rs55635013 | T | A | 3 | 5.51E-06 | 5.51E-06 | 0.7296 | 0.7296 | 0.5619 | 0 | 0.068593489 | Cit-C1(Cit-R) |
| 5 | 72521285 | rs61341658 | C | G | 3 | 7.84E-06 | 7.84E-06 | 0.7337 | 0.7337 | 0.5638 | 0 | 0.070518994 | Cit-C1(Cit-R) |
| 3 | 173372161 | rs4450850 | A | C | 3 | 8.72E-06 | 0.0001343 | 1.4257 | 1.4505 | 0.2727 | 23.05 | 0.072930897 | Cit-C1(R-Cit) |
| 6 | 3250453 | rs12206382 | A | G | 3 | 2.21E-06 | 6.27E-06 | 0.827 | 0.827 | 0.3346 | 8.67 | 0.066780098 | Cit-F4(Cit-Cit) |
| 6 | 3266596 | rs9501985 | A | G | 3 | 1.51E-06 | 0.0005333 | 0.8241 | 0.8252 | 0.1517 | 46.98 | 0.066780098 | Cit-F4(Cit-Cit) |
| 8 | 99452453 | rs6468633 | G | A | 3 | 1.11E-05 | 1.11E-05 | 1.2385 | 1.2385 | 0.4112 | 0 | 0.081865109 | Cit-F4(Cit-Cit) |
| 9 | 4699784 | rs1048020 | G | A | 3 | 3.68E-06 | 3.68E-06 | 0.7903 | 0.7903 | 0.4331 | 0 | 0.066780098 | Cit-F4(Cit-Cit) |
| 1 | 114101039 | rs2359167 | A | G | 3 | 6.82E-06 | 2.19E-05 | 1.2078 | 1.208 | 0.3247 | 11.11 | 0.070518994 | Cit-Vim2-17 |
| 1 | 114102858 | rs61817589 | A | G | 3 | 1.50E-05 | 1.50E-05 | 1.1986 | 1.1986 | 0.3982 | 0 | 0.098660569 | Cit-Vim2-17 |
| 1 | 114105331 | rs6679677 | A | C | 3 | 4.05E-07 | 4.05E-07 | 1.2908 | 1.2908 | 0.5279 | 0 | 0.049374952 | Cit-Vim2-17 |
| 1 | 114113958 | rs4145859 | G | C | 3 | 7.29E-06 | 2.93E-05 | 1.2068 | 1.2071 | 0.3157 | 13.28 | 0.070518994 | Cit-Vim2-17 |
| 1 | 114117743 | rs12041547 | G | C | 3 | 8.25E-06 | 6.17E-05 | 1.2054 | 1.2058 | 0.2891 | 19.43 | 0.071454878 | Cit-Vim2-17 |
| 1 | 114159273 | rs1217413 | G | A | 3 | 1.10E-05 | 7.77E-05 | 1.2024 | 1.2028 | 0.2891 | 19.42 | 0.081728937 | Cit-Vim2-17 |
| 1 | 114179091 | rs2476601 | A | G | 3 | 4.55E-07 | 4.55E-07 | 1.2893 | 1.2893 | 0.5109 | 0 | 0.049374952 | Cit-Vim2-17 |
| 1 | 114216891 | rs2488457 | G | C | 3 | 2.09E-06 | 5.77E-06 | 1.2229 | 1.2232 | 0.3339 | 8.84 | 0.066780098 | Cit-Vim2-17 |
| 1 | 114221851 | rs12566340 | A | G | 3 | 7.03E-06 | 2.67E-05 | 1.2059 | 1.2063 | 0.3173 | 12.87 | 0.070518994 | Cit-Vim2-17 |
| 1 | 114221985 | rs7529353 | A | G | 3 | 7.47E-06 | 2.11E-05 | 1.2051 | 1.2054 | 0.3288 | 10.1 | 0.070518994 | Cit-Vim2-17 |
| 1 | 114222836 | rs11102691 | A | G | 3 | 1.12E-05 | 5.35E-05 | 1.2025 | 1.2031 | 0.3049 | 15.81 | 0.081865109 | Cit-Vim2-17 |
| 1 | 113987113 | rs1230661 | A | G | 3 | 1.04E-05 | 1.04E-05 | 1.2 | 1.2 | 0.5774 | 0 | 0.079750745 | Cit-Vim60-75 |
| 1 | 114101039 | rs2359167 | A | G | 3 | 4.30E-06 | 4.30E-06 | 1.2097 | 1.2097 | 0.5535 | 0 | 0.066780098 | Cit-Vim60-75 |
| 1 | 114102858 | rs61817589 | A | G | 3 | 7.20E-06 | 7.20E-06 | 1.2036 | 1.2036 | 0.5027 | 0 | 0.070518994 | Cit-Vim60-75 |
| 1 | 114105331 | rs6679677 | A | C | 3 | 3.36E-09 | 3.36E-09 | 1.348 | 1.348 | 0.5241 | 0 | 0.002526848 | Cit-Vim60-75 |
| 1 | 114113958 | rs4145859 | G | C | 3 | 3.42E-06 | 3.42E-06 | 1.2118 | 1.2118 | 0.5624 | 0 | 0.066780098 | Cit-Vim60-75 |
| 1 | 114117743 | rs12041547 | G | C | 3 | 3.91E-06 | 3.91E-06 | 1.2104 | 1.2104 | 0.5321 | 0 | 0.066780098 | Cit-Vim60-75 |
| 1 | 114159273 | rs1217413 | G | A | 3 | 4.37E-06 | 4.37E-06 | 1.2093 | 1.2093 | 0.5534 | 0 | 0.066780098 | Cit-Vim60-75 |
| 1 | 114179091 | rs2476601 | A | G | 3 | 2.24E-09 | 2.24E-09 | 1.3526 | 1.3526 | 0.4719 | 0 | 0.002526848 | Cit-Vim60-75 |
| 1 | 114216891 | rs2488457 | G | C | 3 | 2.34E-06 | 2.34E-06 | 1.2188 | 1.2188 | 0.8237 | 0 | 0.066780098 | Cit-Vim60-75 |
| 1 | 114221851 | rs12566340 | A | G | 3 | 3.21E-06 | 3.21E-06 | 1.2111 | 1.2111 | 0.6632 | 0 | 0.066780098 | Cit-Vim60-75 |
| 1 | 114221985 | rs7529353 | A | G | 3 | 4.29E-06 | 4.29E-06 | 1.208 | 1.208 | 0.6402 | 0 | 0.066780098 | Cit-Vim60-75 |
| 1 | 114222836 | rs11102691 | A | G | 3 | 5.04E-06 | 5.04E-06 | 1.2082 | 1.2082 | 0.9332 | 0 | 0.066780098 | Cit-Vim60-75 |
| 1 | 114249912 | rs11552449 | A | G | 3 | 1.40E-05 | 1.40E-05 | 1.2156 | 1.2156 | 0.845 | 0 | 0.094721618 | Cit-Vim60-75 |
| 1 | 114251352 | rs11102701 | T | A | 3 | 7.71E-06 | 7.71E-06 | 1.2236 | 1.2236 | 0.8671 | 0 | 0.070518994 | Cit-Vim60-75 |
| 1 | 114273106 | rs3811019 | C | G | 3 | 9.93E-06 | 0.02231 | 1.3092 | 1.303 | 0.0283 | 71.95 | 0.077078447 | Cit-Vim60-75 |
| 1 | 114276873 | rs17032011 | A | G | 3 | 9.50E-06 | 0.008944 | 1.3079 | 1.305 | 0.0624 | 63.96 | 0.075682517 | Cit-Vim60-75 |
| 2 | 100955499 | rs6757524 | G | A | 3 | 1.18E-05 | 0.0004932 | 1.1845 | 1.1835 | 0.2099 | 35.94 | 0.084712455 | Cit-Vim60-75 |
| 25 | 1879902 | rs5989956 | G | A | 3 | 1.42E-05 | 1.42E-05 | 1.1903 | 1.1903 | 0.7161 | 0 | 0.095489802 | Cit-Vim60-75 |
